# Effect of discussing personalized estimates of diabetes risk for people with prediabetes

**DOI:** 10.1101/2025.10.02.25337167

**Authors:** Natalia Olchanski, Christine E. Skiro, Kristen Sonon, Sarah Kaus, Ericka C. Holmstrand, Elizabeth L. Ciemins, John K. Cuddeback, Francis R. Colangelo, David M. Kent

## Abstract

**Background:** To assess acceptability, feasibility, and effectiveness of incorporating individualized risk prediction into clinical assessment, decision making, and communication of risk of type 2 diabetes, with and without preventive interventions, in patients with prediabetes.

**Methods:** We integrated a prediction model into the clinical workflow at a US health care organization. We conducted patient and provider focus groups and pre- and post-dissemination surveys among 2,500 patients with prediabetes who had primary care visits between May 2018 and December 2019. We compared rates of progression to type 2 diabetes at 3 years between the intervention group and a propensity score matched cohort of patients who received usual care.

**Results:** Prior to implementing the predictive model, 41.6% of providers and 63.8% of patients felt confident or very confident in their ability to estimate the risk of progression to diabetes for individual patients. After personalized risk information was made available, this increased to 92.8% and 66.9%, for providers and patients, respectively. People with prediabetes who had a discussion with their provider about their personal risk of developing type 2 diabetes, supported by the EHR-based prediction model, were significantly less likely to progress to diabetes in the following 3 years, compared to a propensity-score-matched cohort who received usual care in the same health system without individualized risk estimates (19.5% vs. 27.6%, p = 0.042).

**Conclusions:** Used at the point of care during a shared decision-making discussion between the patient and provider, the EHR-based diabetes risk calculator helped providers prioritize patients for diabetes prevention interventions, facilitated communication, and improved health outcomes among patients with prediabetes.

**Contributions to the Literature:**

- One-third of U.S. adults have prediabetes, a condition which carries an increased risk of developing type 2 diabetes, yet few people with prediabetes receive evidence-based preventive intervention such as the Diabetes Prevention Program (DPP) or metformin. A recent study suggests a new category of preventive intervention, GLP-1 and GIP receptor agonist, is highly effective for prevention in people with prediabetes and obesity. Provider organizations need an efficient way to risk-stratify the large population of people with prediabetes, in order to focus prevention efforts toward those who are at greatest risk.
- A risk calculator integrated in the electronic health record (EHR) equipped physicians to provide an estimate of each patient’s individual risk of developing type 2 diabetes and prioritize patients for preventive interventions. DPP referrals and metformin prescribing increased substantially, and patients at high risk were more likely to receive a preventive intervention than those at intermediate or low risk.
- People with prediabetes for whom use of the diabetes risk prediction tool was documented were significantly less likely to progress to diabetes over 3 years, compared to a propensity-score-matched cohort who received usual care in the same health system without individualized risk estimates (19.5% vs. 27.6%).

## Background

Prediabetes is highly prevalent, affecting more than one-third of U.S. adults.(1, 2) While everyone with prediabetes is at increased risk of developing type 2 diabetes, an individual’s risk depends on many factors and has been shown to vary widely.(3) In the Diabetes Prevention Program (DPP) Study,(4–6) two interventions were shown to reduce the risk of developing diabetes among people with prediabetes—an intensive lifestyle program and, to a lesser extent, taking metformin. Recently, the SURMOUNT-1 trial added a third evidence-based intervention, showing that taking tirzepatide dramatically reduces the risk of developing diabetes among people with prediabetes and obesity (hazard ratio 0.07 at 40 months, P < 0.001).(7) Although these interventions are validated, there are drawbacks. The DPP requires enrollment in a structured program with limited capacity and considerable effort of the participant,(8, 9) and tirzepatide is expensive and not without side effects. Practically, provider organizations need ways to risk-stratify the large population of people with prediabetes so they can focus the care team’s effort and payment for interventions on those patients who are likely to gain the greatest benefit, i.e. those at higher baseline risk of developing diabetes.(3)

Our team has previously developed clinical prediction models to estimate the risk of developing diabetes in patients with prediabetes and project the risk-specific effects of the DPP lifestyle intervention and/or metformin, using both observational and randomized data.(10)

We report here two new studies. First, an implementation study focused on the feasibility and value of incorporating individualized risk prediction into clinical encounters in the primary care offices of a multi-specialty medical group in the U.S. The effect on referral for the DPP and prescribing metformin was also assessed. Second, an outcome study followed a subset of patients from the first study to compare their likelihood of developing diabetes within 3 years against a matched control population who received usual care in the same health system without individualized risk estimates.

## Methods

### Implementation Study

#### Study Setting and Population

We conducted a study to assess the use of individualized risk prediction in clinical assessment, communicating the risk of developing type 2 diabetes and recommendations for prevention, and decision-making at Premier Medical Associates (PMA), a 100-provider multi-specialty medical group in suburban Pittsburgh, PA. The study included patients who had primary care visits from May 1, 2018, through December 31, 2019, were aged 18–75 at the time of the visit, and had evidence of prediabetes: no diagnosis of diabetes on their problem list in the electronic health record (EHR), and their last HbA1c within a year prior to the visit was in the range 5.7–6.4% (39–47 mmol/mol).

#### Risk Prediction Model

In an earlier study funded by the Patient-Centered Outcomes Research Institute (PCORI), (10) our team at the Predictive Analytics and Comparative Effectiveness (PACE) Center at Tufts Medical Center and the American Medical Group Association (AMGA) developed an EHR-based risk prediction model that provides individualized estimates to support targeted prevention for people with prediabetes. It calculates their risk of developing type 2 diabetes within 3 years and the benefit they would experience from the interventions evaluated in a landmark clinical trial, the DPP Study—either an intensive lifestyle program or taking metformin.(4–6) We developed and validated the risk model using data from 2.2 million people with prediabetes in the Optum Labs Data Warehouse (OLDW), a de-identified database of healthcare claims and clinical data containing longitudinal health information on enrollees and patients, representing a mixture of ages and geographic regions across the U.S. The model uses 11 variables that are typically available in an EHR (Table 1) and accommodates missing data. We used data from 3,081 participants in the DPP Study(4) to estimate risk-specific treatment effects.

**Table 1.** EHR data elements included as predictors to estimate the risk of developing type 2 diabetes within 3 years, for people with prediabetes (Kent et al.(10))

|  |
| --- |
| Age |
| Sex |
| Race (Asian, Black, White, Other)* |
| Smoking status (Current, Former, Never)* |
| Hypertension diagnosis (Yes, No) |
| Systolic blood pressure* |
| Hemoglobin A1c* |
| Fasting plasma glucose* |
| Triglycerides* |
| HDL cholesterol* |
| Body mass index* |

\* Model is designed to make predictions even when data are missing for one or more of the parameters with an asterisk.

The implementation study focused on use of this model at the point of care for people with prediabetes in PMA’s 7 primary care offices, as part of an informal shared decision-making conversation. The risk prediction model was implemented in PMA’s Allscripts TouchWorks EHR, using Galen eCalcs, which retrieved the model parameters from the patient’s file and displayed them for review and editing by the clinician. The clinician could run the calculator during a visit, but in most cases, a member of the care team ran the calculator the day before a scheduled visit, and the results were discussed during the morning huddle before the visit.

#### Implementation Study Measures

The study began with provider and patient focus groups to inform implementation, specifically how the patient- and provider-facing tools would look and be used in practice. Separate focus group guides were developed for the provider and patient cohorts. Providers were recruited from the pool of primary care practices that would be using the risk calculator. Patient focus groups were drawn from organizational patient councils and patients referred by project team members. These sessions explored patients’ understanding and acceptance of the risk calculator and how best to communicate risk estimates. Focus groups were audio recorded and transcribed and results were analyzed using a rapid qualitative analysis approach informed by the framework method which uses a matrix format to identify common themes.(11, 12)

Pre- and post-implementation surveys were conducted with all PMA primary care providers and separate random samples of patients, drawn from all patients with evidence of prediabetes who were seen at PMA for well visits. Survey questions addressed process and experience of both understanding the risk of developing diabetes and making decisions about preventative care. Providers at PMA received different pre- and post-surveys through email links to SurveyMonkey. Patients received the same 12-item questionnaire, pre- and post-, through standard mail, with a study information sheet, a $2 bill as compensation, and an addressed, stamped return envelope. Survey responses were stored and processed without linking to respondent identity, to protect participant anonymity and confidentiality. Survey responses before and after implementation were compared using chi-square tests for homogeneity of proportions. Questionnaires and detailed survey results are included in Additional File 1. The consensus-based checklist for reporting of survey studies (CROSS) is in Additional File 2.

Implementation included preparation meetings with providers and care teams in each PMA primary care office that focused on the need addressed by the model, use of the calculator in the EHR, and how to interpret the results and explain them to patients. Data extracted from the EHR were used to track use of the risk calculator by providers and outcomes of informal shared decision-making conversations about patients’ personal risk estimates (e.g., if patients received referral for the year-long DPP lifestyle program or were prescribed metformin).

The study received IRB approval from Tufts Medical Center.

#### Outcome Study

The outcome study assessed the impact of using personalized diabetes risk prediction, as implemented at PMA, on developing type 2 diabetes within 3 years. The intervention group was drawn from the PMA patients in the implementation study described above who were insured by one of Highmark Health’s companies, which covered approximately 45% of the PMA patient population during the study period. PMA is a part of the Allegheny Health Network (AHN), but at the time of the project implementation AHN’s clinicians were using a different EHR and did not have the risk model available to guide patient care. We used propensity score matching(13, 14) to draw a control group of similar patients seen elsewhere within AHN. Both propensity score matching and ascertainment of developing type 2 diabetes used EHR data from PMA and AHN, as well as adjudicated claims data and case management data from Highmark.

The study included adult patients with at least 4 years of continuous insurance enrollment in a commercial or Medicare Advantage (MA) plan with Highmark that included prescription drug coverage. The control sample was drawn from patients with prediabetes who were seen for primary care visits at AHN (scheduled at least 5 days in advance and completed between May 1, 2018, and December 31, 2019) and also had an HbA1c result in the prediabetes range (5.7–6.4%) within 2 years prior to the visit. The study excluded patients with any evidence of diabetes before the index primary care visit—diagnosis of type 1 or type 2 diabetes in medical claims, drug claims for an antihyperglycemic drug (see Additional File 3), enrollment in a diabetes support program, or laboratory evidence of diabetes (HbA1c). Additional exclusion criteria were hospice care enrollment status, missing data relating to the propensity score model, or conflicting data relating to parameters of the risk model.

Separate propensity score models were developed for the commercially insured and MA populations. Both models included the same 15 parameters (Table 2), reflecting demographics, claims-based risk scores, healthcare utilization, clinical data (including predicted diabetes risk using this risk model), as well as receptivity to and enrollment in Highmark case management programs. Matched controls were selected separately for the commercial and MA intervention populations, then combined for the analysis of progression to diabetes. We matched the cohorts using PSMATCH procedure in SAS software utilizing greedy matching. In the propensity score-matched sample, balance between groups was assessed using standardized differences, with values <0.2 considered negligible. Consistent with recommendations that hypothesis testing is not appropriate for baseline characteristics, p-values are not reported.(15, 16)

**Table 2.** Parameters used in the propensity score models for creating the matched control population.

|  |  |
| --- | --- |
| Demographics | Age |
|  | Gender |
| Risk Scores | Charlson Comorbidity Index |
|  | Prospective risk score (Highmark internal model) |
|  | Care gap index (Cotiviti) |
|  | High-cost claims score (Cotiviti) |
|  | Likelihood of emergency dept. visit (Cotiviti) |
|  | Likelihood of hospitalization (Cotiviti) |
| Utilization | PCP visits (during 12 months before initial primary care visit) |
|  | Specialist visits |
| Clinical | Predicted diabetes risk (using this model) |
|  | Body mass index |
|  | Hyperlipidemia (Cotiviti criteria) |
| Chronic Care Programs | Receptivity to enrolling in a program (Highmark internal model) |
|  | Enrolled chronic care programs (pre- period) |

Cotiviti risk scores and chronic condition flags are based on medical claims data. The algorithms are proprietary, but these risk scores are widely used within the industry.

For the outcome study, the intervention was simply receiving a personalized estimate of diabetes risk as part of a discussion with one’s provider, and the outcome was development of type 2 diabetes within 3 years following the index primary care visit, without regard to any specific recommendation by the provider or any specific action by the patient. Development of type 2 diabetes was defined as (a) 2 or more medical claims with a diagnosis of type 2 diabetes (E11); (b) at least 1 claim with a diagnosis of type 2 diabetes plus prescription drug claim(s) for at least 60 days supply of a diabetes medication (see Additional File 3); and/or (c) HbA1c ≥ 6.5% indicated in the EHR. Rates of development of diabetes at 3 years were compared using McNemar’s test.

This study was determined exempt by the AHN Institutional Review Board.

## Results

### Implementation Study

#### Patient and Provider Focus Groups (pre-implementation)

Provider focus groups included 7 to 11 primary care team members. Patient focus groups consisted of 6 to 8 patients. Consistently, people with prediabetes said they wanted a personalized estimate of their risk of developing diabetes. In each patient focus group, at least one participant spontaneously quoted the ages at which several family members had developed type 2 diabetes; once the topic had been raised, most participants quoted ages within their own families. This suggests that they were already thinking about their personal risk in probabilistic terms, using the information they had available. Similarly, providers said they wanted individualized risk estimates for their patients, both to inform shared decision-making and to aid in prioritization of intervention for people with prediabetes. Providers felt they could give only limited, generalized guidance to their patients with prediabetes. For example, they could say that prediabetes carries an increased risk of developing diabetes, but they were unable to offer a quantitative estimate, which patients often requested.

In the focus groups, providers and patients alike perceived specific estimates of both baseline risk of diabetes and the potential benefits of preventive interventions as helpful for clear understanding, communication, and decision-making. Patients emphasized the importance of information sharing and coordination among members of their care teams, and they felt that documentation of individualized risk estimates would facilitate this. Importantly, patients said they would regard knowing that their personal risk was high, compared to other people with prediabetes, as a strong motivator for behavior change.

#### Pre- and Post-Implementation Surveys of Providers and Patients

Survey responses at PMA included 162 patients (24% response rate) and 24 providers (71% response rate) for pre-implementation and 171 patients (30% response rate) and 28 providers (82% response rate) for post-implementation surveys.

Without the model (pre-implementation), 41.6% of providers felt confident or very confident in their ability to estimate the risk of progression to diabetes for individual patients, but with the model, this increased to 92.8% (p<0.001, Figure 1). Patients demonstrated higher baseline confidence compared to providers, but the increase after implementation of the diabetes risk estimation model was not statistically significant (63.8% vs. 66.9%, p=NS; Figure 1).

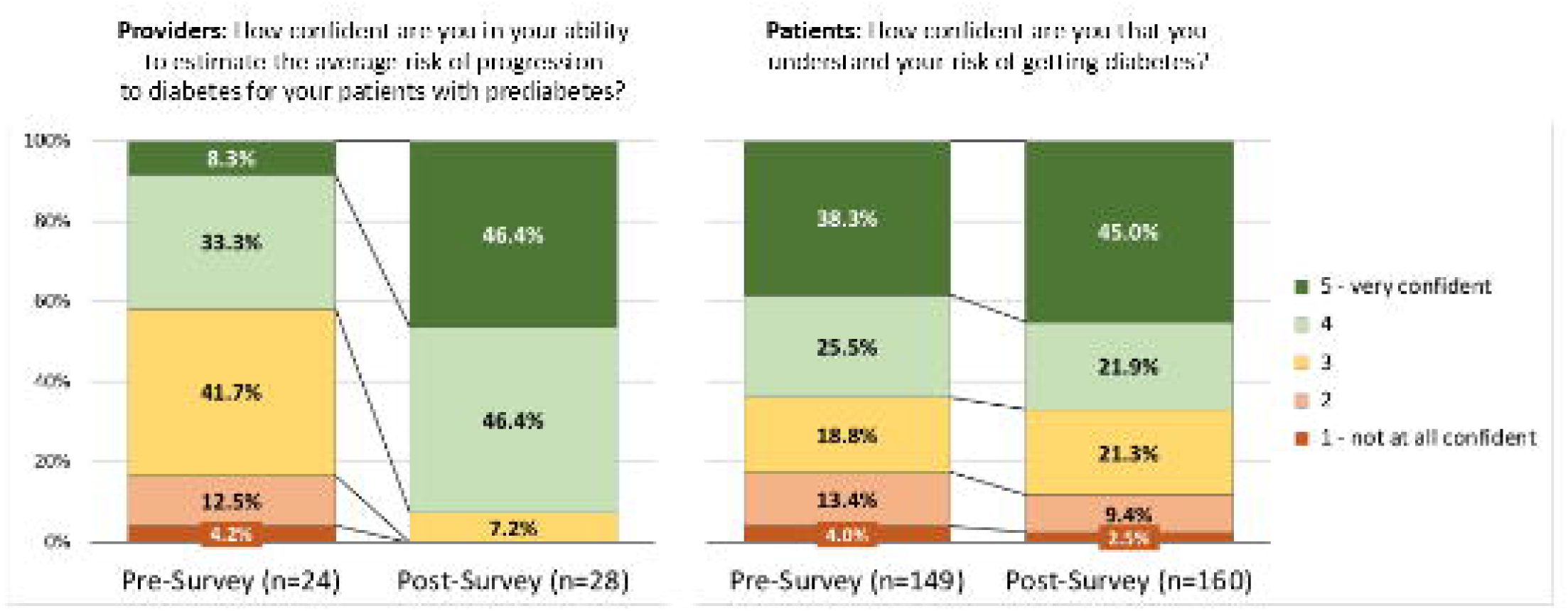

With the model’s individualized estimates of the benefits of the DPP and metformin, providers felt more confident about tailoring their recommendations for diabetes prevention (45.9% felt confident or very confident without the model vs. 96.4% with the model, p<0.001; Figure 2) and about communicating the benefit that each patient could anticipate (54.2% vs. 100%, p<0.001; Figure 2).

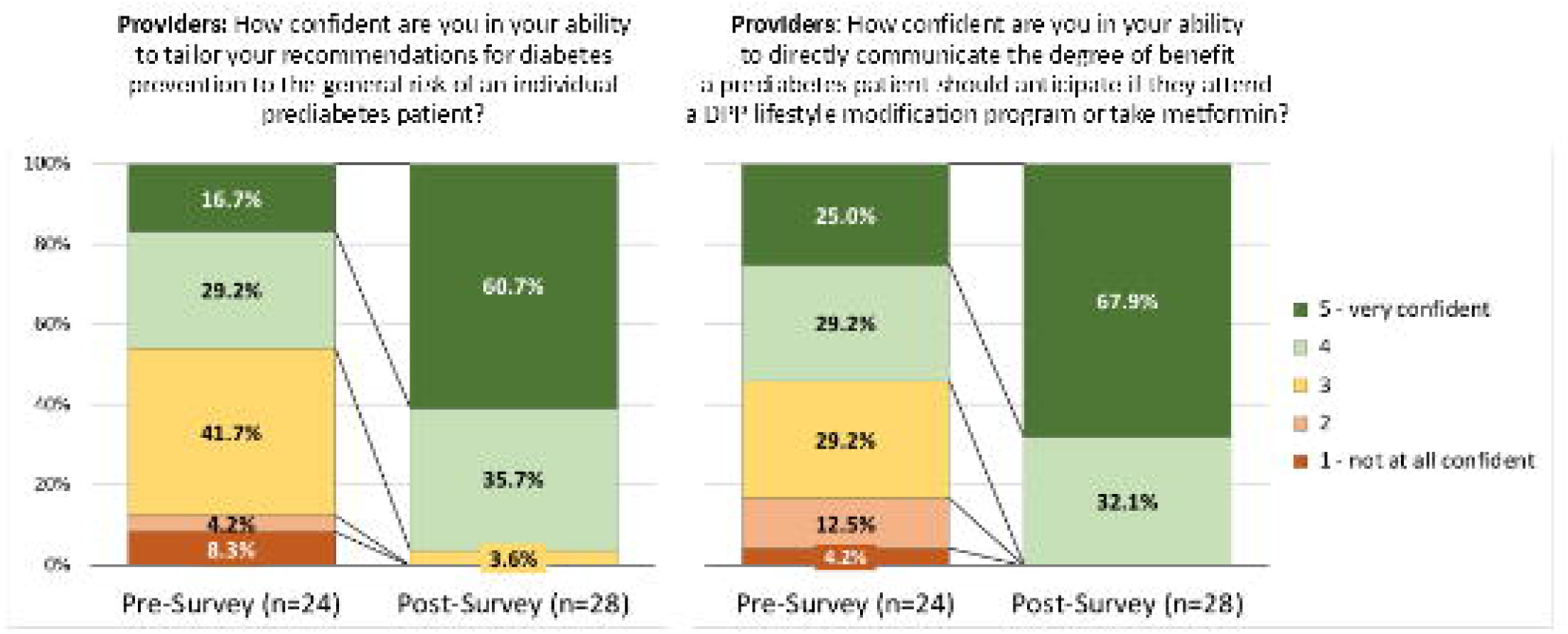

In both the pre- and post-implementation surveys, patients’ most preferred source of information about their risk of diabetes was talking to their provider (85.9% and 87.0% for talking with their provider, pre- and post-, vs. 32.9% and 27.2% for their next-most preferred source, reading materials; Figure 3). When the risk model was used by their provider during an office visit, more patients said the conversation with the provider helped them understand their risk of developing diabetes (32.8% in the pre-survey vs. 46.8% in the post-survey, p=0.003), but their overall satisfaction with the conversation with their provider about their risk of diabetes was quite similar, pre- and post-implementation (Figure 4).

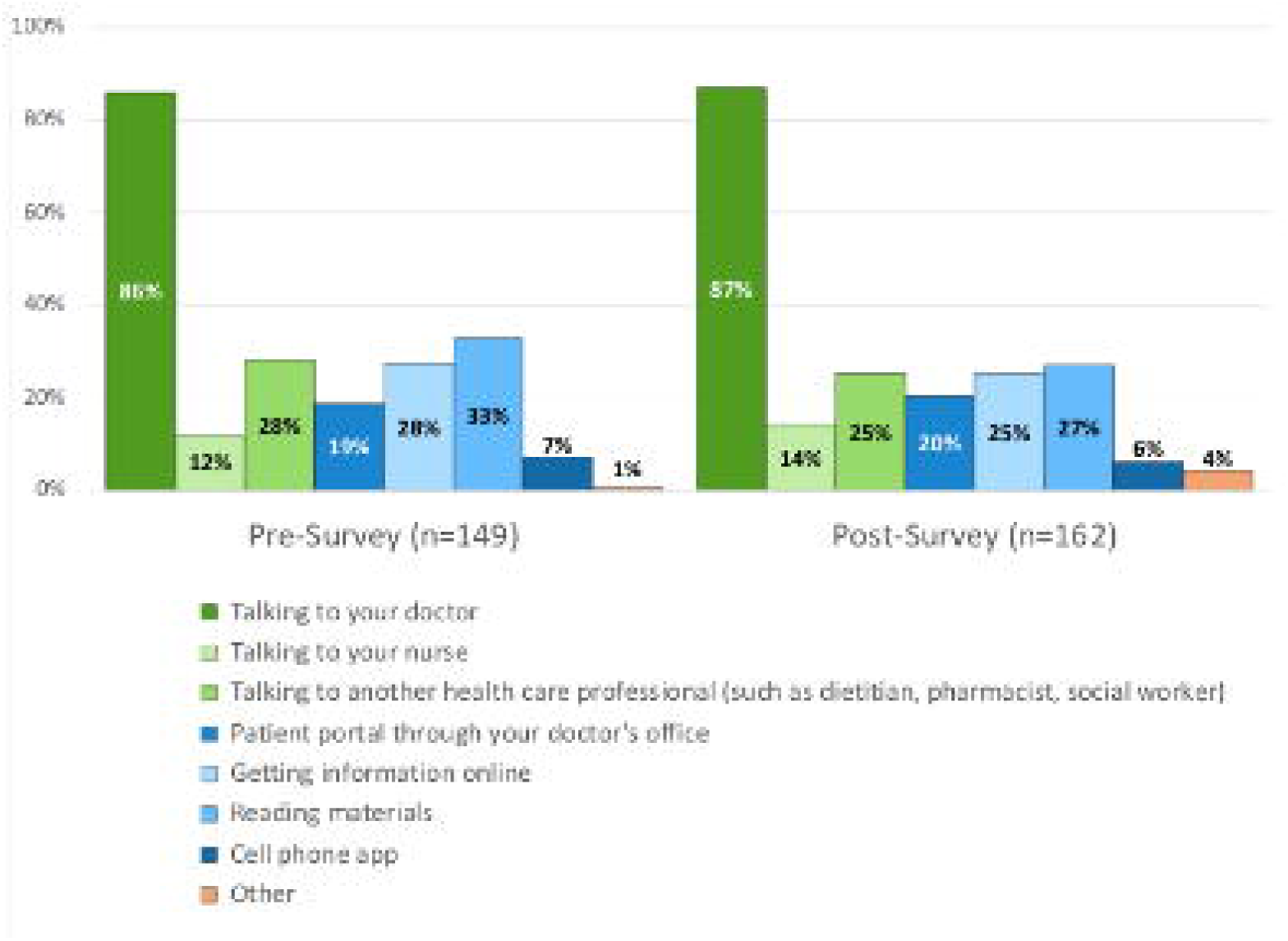

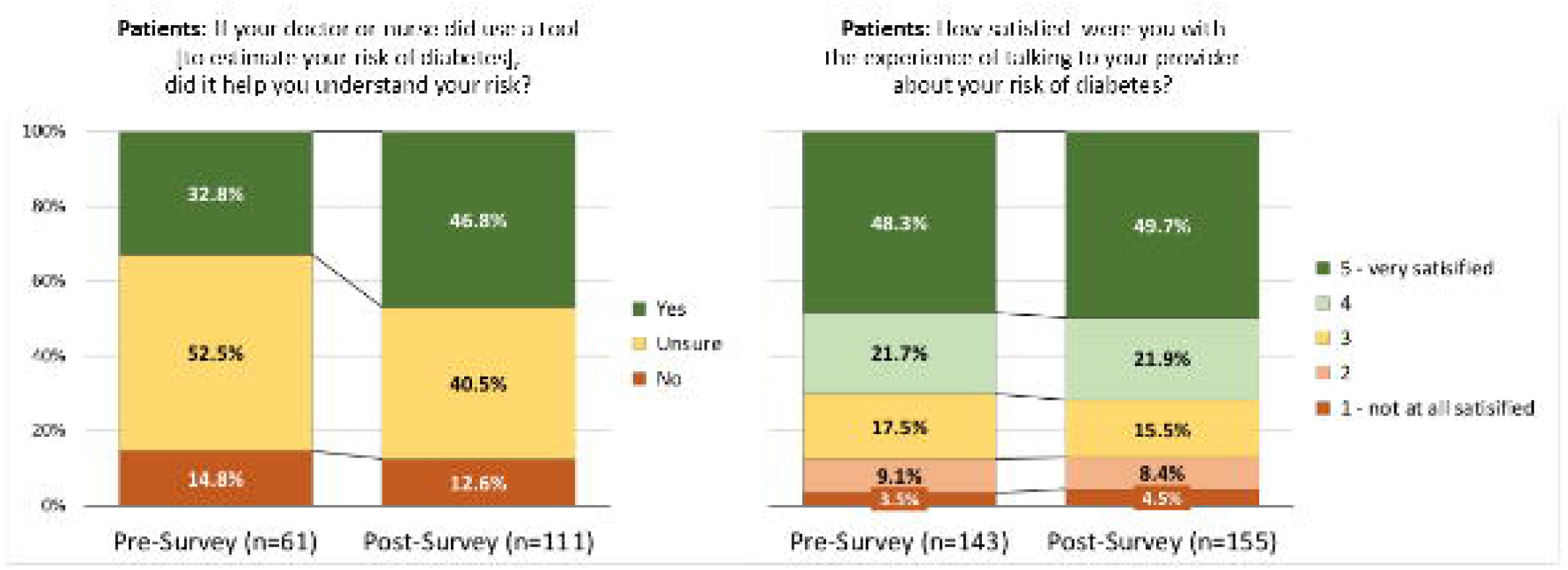

#### Use of the Model by Providers and Preventive Interventions

From May 2018 to December 2019, about 2,500 patients with prediabetes were seen in primary care at PMA. Within the first 9 months, the model had been run at least once for 52% of these patients; by 20 months, risk had been calculated for 82% (Table 3). Providers became increasingly comfortable with the model over time, anecdotally and as indicated by the increase in reported confidence from the pre- to post-surveys (Figure 2).

**Table 3.** Rates of risk calculator usage over time at Premier Medical Associates during the study period.

|  | 5/1/18–<br>1/31/19<br>(9 months) | 5/1/18–<br>5/31/19<br>(13 months) | 5/1/18–<br>8/31/19<br>(16 months) | 5/1/18–<br>12/31/19<br>(20 months) |
| --- | --- | --- | --- | --- |
| People with Prediabetes | 2,466 | 2,425 | 2,518 | 2,454 |
| Calculation Completed | 1,272 | 1,566 | 1,881 | 2,023 |
| Percent with Calculation | 52% | 65% | 75% | 82% |

Patient counts in each column reflect all patients with prediabetes, aged 18–75 at their first visit within the time period, whose last HbA1c during the time period was in the range 5.7–6.4%, who had at least one visit with a primary care provider during the time period, and who were still an active patient of PMA at the end of the time period.

Before the model was implemented at PMA, no patient with prediabetes had been formally referred to the DPP at the local YMCA (although 5% of patients in the pre-survey reported that their provider had recommended the DPP), and fewer than 5% of people with prediabetes had been prescribed metformin. Following implementation of the risk prediction model, including the provider education and discussion involved with its implementation, preventive interventions increased greatly and became strongly risk-stratified, with 74% of high-risk patients (top quartile of risk scores), 19% of intermediate-risk patients (middle half), and 8% of low-risk patients (bottom quartile) receiving an intervention, such as referral to DPP or prescription of metformin. Among high-risk patients receiving an intervention, 71% were referred to the DPP, and 29% were prescribed metformin.

#### Patient Follow-through on Referrals to the DPP

Of 487 people with prediabetes who were referred to the DPP in the first three months after the model was implemented at PMA, 124 (25%) called the YMCA to inquire, and 64 (13%) enrolled in the DPP. The PMA patients who completed the year-long DPP program achieved an average weight loss of 7.4% (the “design goal” of the DPP is 7%).

### Outcome Study

#### Characteristics of the Study Sample

After applying inclusion and exclusion criteria, the intervention group included 148 commercially insured and 120 MA patients at PMA, and the pool of potential controls included 1,403 commercially insured and 741 MA patients seen elsewhere at AHN, where the risk model was not available. After propensity score matching, the intervention group and control group each had 124 commercially insured and 97 MA patients (Additional File 3). The intervention and control groups each contained 221 patients with an average age of 62.1 and 62.4 years, respectively. The groups were each 56.6% female, with average predicted risks of progression to diabetes of 35% (Table 4). The most common comorbidities were hypertension (61.5% of the intervention group and 67.4% of the control group) and hyperlipidemia (54.3% of the intervention group and 59.7% of controls). The intervention and control groups were well matched on several risk scores, including Highmark’s prospective risk score, Cotiviti measures (likelihood of hospitalization, likelihood of emergency department visit, and care gap index), BMI, and receptivity to, and participation in, chronic care programs. The groups had similar rates of comorbidities, with slightly higher rates of hypertension and hyperlipidemia in the control group (see above), a higher rate of malignant neoplasm in the control group (4.1% intervention vs. 7.7% control), and a higher rate of depression in the intervention group (11.8% intervention vs. 8.6% control). The control group had a higher proportion living in a low-income census tract (19.0% intervention vs. 29.0% control), but similar proportions reported living alone (46.6% intervention vs. 43.9% control).

**Table 4.** Population characteristics for outcome study, comparing patients with prediabetes who received individualized diabetes risk prediction (intervention) against a propensity-score-matched cohort who received usual care, without personalized prediction, within the same health system (control).

|  | **Intervention**<br>**(N=221)** |  | **Control**<br>**(N=221)** |  |  |
| --- | --- | --- | --- | --- | --- |
|  | **mean**<br>**or %** | **SD** | **mean**<br>**or %** | **SD** | **Std.**<br>**Diff.** |
| Demographics |  |  |  |  |  |
| Age | 62.1 | 8.84 | 62.4 | 9.09 | -4.2% |
| Gender (female) | 56.6% |  | 56.6% |  | 0.0% |
| Risk Scores |  |  |  |  |  |
| Charlson Comorbidity Index | 0.67 | 1.28 | 0.61 | 1.00 | 5.9% |
| Prospective risk score (Highmark model) | 1.42 | 2.01 | 1.51 | 2.16 | -4.6% |
| Care gap index (Cotiviti) | 2.23 | 1.97 | 2.33 | 2.03 | -5.0% |
| High-cost claims score (Cotiviti) | 1.29 | 1.44 | 1.37 | 1.40 | -5.6% |
| Likelihood of emergency dept. visit (Cotiviti) | 0.10 | 0.05 | 0.09 | 0.05 | 8.9% |
| Likelihood of hospitalization (Cotiviti) | 0.05 | 0.05 | 0.05 | 0.03 | 8.2% |
| Chronic Care Programs |  |  |  |  |  |
| Receptivity to enrolling (Highmark model) | 0.34 | 0.11 | 0.35 | 0.12 | -5.8% |
| Number of enrolled program (pre- period) | 0.22 | 0.58 | 0.20 | 0.51 | 2.5% |
| Clinical |  |  |  |  |  |
| Body mass index (BMI) | 31.9 | 7.1 | 31.4 | 9.7 | 5.1% |
| Predicted diabetes risk (using study model) | 0.35 | 0.11 | 0.35 | 0.10 | 0.3% |
| Comorbidities (prior to initial primary care visit)* |  |  |  |  |  |
| Hypertension | 61.5% |  | 67.4% |  | -12.3% |
| Hyperlipidemia | 54.3% |  | 59.7% |  | -11.0% |
| Coronary artery disease | 9.1% |  | 7.2% |  | 6.6% |
| Congestive heart failure | 2.3% |  | 2.7% |  | -2.9% |
| Chronic kidney disease | 2.7% |  | 3.2% |  | -2.7% |
| Asthma | 5.0% |  | 5.0% |  | 0.0% |
| Chronic obstructive pulmonary disease | 6.8% |  | 6.3% |  | 1.8% |
| Depression | 11.8% |  | 8.6% |  | 10.5% |
| Other behavioral health condition | 15.4% |  | 16.3% |  | -2.5% |
| Substance use disorder | 5.4% |  | 5.9% |  | -2.0% |
| Malignant neoplasm | 4.1% |  | 7.7% |  | -15.4% |
| Social determinants of health (Highmark model) |  |  |  |  |  |
| Has financial stability claim | 0.0% |  | 0.5% |  | — |
| Has social stability claim | 0.5% |  | 2.3% |  | -15.7% |
| Has social support claims | 4.5% |  | 1.8% |  | 15.6% |
| Lives in low-income census tract | 19.0% |  | 29.0% |  | -23.5% |
| Lives alone | 46.6% |  | 43.9% |  | 5.5% |

Additional details for Predicted diabetes risk (using study model):
Intervention group – mean 0.35, median 0.35, minimum 0.12, maximum 0.61
Control group – mean 0.35, median 0.33, minimum 0.15, maximum 0.63
\* Comorbidities determined by Cotiviti condition categories.

#### Effect of Intervention on Development of Diabetes

Among the intervention group, 19.5% progressed to diabetes within 3 years, compared to 27.6% in the control group, p = 0.042. For 74% of these individuals, development of diabetes was ascertained using 2 or more medical claims with a diagnosis of type 2 diabetes, while 7% had a diagnosis on 1 medical claim plus at least 60 days supply of a diabetes medication in pharmacy claims. For 18.6% of those who developed diabetes in the intervention group and 19.7% in the control group, there was no diagnosis on a claim, but there was an HbA1c ≥ 6.5% in their EHR data (Table 5).

**Table 5.** Diabetes progression at 3 years in patients with prediabetes who received individualized diabetes risk prediction (intervention) compared to a propensity-score-matched cohort who received usual care, without personalized prediction, within the same health system (control).

|  | Intervention (n=221) |  | Control (n=221) |  |  |
| --- | --- | --- | --- | --- | --- |
|  | n | % | n | % | p |
| 2+ claims with type 2 diabetes Dx | 32 | 14.5% | 45 | 20.4% |  |
| Combined Dx/Rx criteria | 3 | 1.4% | 4 | 1.8% |  |
| HbA1c ≥ 6.5* | 8 | 3.6% | 12 | 5.4% | |
| **Total** | **43** | **19.5%** | **61** | **27.6%** | **0.042** |

\* HbA1c ≥ 6.5 includes only members without a medical/Rx claim for diabetes.

## Discussion

This study assessed the feasibility and impact of using an individualized risk prediction model to estimate the three-year likelihood of developing type 2 diabetes among people with prediabetes. The implementation study demonstrated that the DPP risk model significantly increased provider confidence in estimating individual patient risk and tailoring diabetes prevention recommendations. Use of the DPP risk tool led to both an increase in the rates at which prevention intervention were recommended and a shift in patient selection to better target the high-risk patients most likely to benefit from intervention, in theory improving the efficiency of care under conditions of limited capacity.(17) Furthermore, the related outcome study showed that exposure to the tool providing individualized risks during an encounter was associated with a significant reduction in the development of type 2 diabetes, compared to a matched control group.

The ability to provide individualized diabetes risk estimates represents a meaningful advance in the management of prediabetes. Clinicians often feel overwhelmed by the large number of patients with prediabetes, with a wide variation in the risk of developing diabetes, making it difficult to prioritize intervention referrals. Capacity of labor intensive DPP programs is often limited, and pharmacological interventions can be costly and incur undesired side effects, highlighting the need to prioritize referrals to those patients most likely to benefit. By integrating a risk prediction model into routine care, healthcare providers can more effectively target interventions to those at greatest risk, thereby improving both clinical outcomes and resource allocation. The findings suggest that even modest interventions, such as a simple conversation about risk, can have tangible benefits, likely because they prompt patients to engage more actively in their own care—even in the absence of protocol-driven intervention.

While the risk model appears to be a useful tool for providers, in terms of improving their confidence, our study also highlights the importance of provider-patient communication in fostering trust and motivating behavior change.

Previous studies, such as those from the Diabetes Prevention Program (DPP), have shown that lifestyle interventions and metformin can significantly reduce the risk of developing type 2 diabetes in people with prediabetes.(4–6) However, these interventions are resource-intensive and often underutilized,(8, 9) particularly as approximately one-third of the adult US population meets criteria for prediabetes and most practices lack the means for precise identification of higher risk individuals.(2) Additionally, with new more expensive pharmacotherapies for prediabetes, such as tirzepatide, being introduced into the market, the need for risk stratification to ensure cost-effective care is increasingly acute. With the current high cost of GLP-1 and GIP receptor agonist medications, payers and provider organizations with risk contracts will need to weigh this expense against the likely clinical benefits, which will depend on each person’s baseline risk. The present study’s results suggest that engaging patients in risk discussions has the potential to shift patient selection of preventative interventions to those at higher risk, improving cost-effectiveness and meaningfully impacting outcomes.

Our risk tool has several strengths that are important to highlight. First, the tool integrated 11 clinical factors to determine risk. This permits more precise targeting of patients at higher risk than the common practice of using HgBA1c alone. For example, in our study based on more than 2 million prediabetic patients in the OptumLabs data warehouse, 15% of people in the lower risk quartile had A1c ≥ 6.0, the high end of the prediabetes range. Conversely, in the highest-risk quartile, more than 25% of people had A1c < 6.0. A second strength of the tool is that it was derived from the DPP randomized study, and therefore provides counterfactually valid individualized estimates of the contrast in risk with different interventions, supporting decision-making. Finally, the tool is EHR compatible and robust to missing data, enhancing the usability of the tool for integration into the clinic workflow.

Our study has several strengths. First, the outcome study relied on a propensity score-matched control group, ensuring that the intervention and control groups were well-matched on key baseline characteristics. Second, the real-world setting of both studies (Premier Medical Associates) provides a strong context for understanding the feasibility and impact of integrating this model into routine care. Finally, the multi-specialty approach involving both providers and patients in focus groups ensured that the implementation was user-centered and relevant to both parties’ needs.

Nevertheless, our study does have some limitations. In particular, we implemented this study within a single network. Successful implementation of this tool may depend on system specific features that might not be universally available, such as having a strong champion to help ensure uptake. The outcome study similarly may be confounded by practice-level differences that were not accounted for in the propensity score or by other unmeasured confounders. An ideal design to establish causality would have a concurrent control group within the same system to improve exchangeability between the intervention and control groups and account for any secular changes, although such a design might be vulnerable to contamination.

How much of the effect on outcomes is due to the conversation about the risk estimate, and how much to the broader focus on prediabetes that came with the implementation study? PMA tracked 3 chronic care “balance” measures through the first 12 months of implementation—rates of pneumococcal vaccination, colorectal cancer screening, and breast cancer screening. Across PMA’s 7 primary care offices, the first 2 measures were within ±2%, and mammography ranged from unchanged to +8%. In fact, the lifestyle discipline of the DPP— and even the broader conversation with patients about prevention—may also benefit other metabolic and cardiovascular conditions, which were by far the most common comorbidities in this population with prediabetes.

Future research is needed to further explore the generalizability of our results to other health care systems, including more diverse populations, and to examine longer term outcomes and link changes in outcomes to the interventions received. Additionally, directly measuring costs would help examine whether the improvements in cost-effectiveness (and potentially cost-savings) that would theoretically be anticipated from better targeting are actually realized.

In conclusion, this study highlights the value of incorporating individualized risk estimates for people with prediabetes into clinical practice. By enabling providers to offer personalized and counterfactually valid diabetes risk assessments, the model helps prioritize interventions and fosters more engaged, informed patient-provider discussions. The outcome study provides evidence that these conversations, even in the absence of protocol-driven follow-up interventions, can significantly reduce the risk of progression to type 2 diabetes. Given the growing burden of prediabetes, integrating personalized risk prediction models into routine care represents a promising strategy to improve patient outcomes and optimize the use of healthcare resources.

## Supporting information

Additional File 1. Implementation Study

Additional File 2. CROSS Checklist.docx

Additional File 3. Outcomes study additional tables and figures

## Declarations

### Ethics approval and consent to participate

The implementation study was approved by the Tufts Health Sciences Institutional Review Board. Informed consent was obtained from participants. The outcome study was determined exempt by the Allegheny Health Network Institutional Review Board.

### Implementation Note

The diabetes risk model developed by Tufts and AMGA(10) is now available as a cloud-hosted “SMART on FHIR” app from Elimu Informatics. (Acronyms: Substitutable Medical Applications and Reusable Technologies, built on the framework of the HL7 Fast Healthcare Interoperability Resources standard.) This app has been implemented within the Epic EHR at Tufts Medical Center.

### Availability of data and material

Deidentified data from the implementation study and a detailed summary of the outcome analysis conducted by Highmark Health are available from the corresponding author upon reasonable request.

## Authors’ contributions

NO, JKC, ELC, and DMK drafted the manuscript. ELC, JKC, and DMK oversaw and designed the study. JKC and DMK obtained funding for the research. FRC led the implementation at PMA. CS, KS, SK, and ECH led the outcome study at Highmark Health. All authors contributed to data access and collection, analysis, development of conclusions, and all authors reviewed and contributed significantly to the final manuscript. All authors approve the final version of the manuscript.

## Acknowledgements

The authors wish to thank Carolyn Koenig, MD, of Mercy, in St. Louis, who led an effort similar to that at PMA which had to be discontinued just prior to implementation due to the COVID-19 pandemic. Pre-implementaiton focus group and survey results at Mercy were very similar to those at PMA.

## Additional Files

### Additional File 1. Implementation Study

Patient pre- and post-implementation questionnaires Provider pre- and post-implementation questionnaires Patient survey responses, pre- and post-implementation Provider survey responses, pre- and post-implementation

### Additional File 2. CROSS Checklist.docx

Consensus-Based Checklist for Reporting of Survey Studies (CROSS) filled out for the study.

### Additional File 3. Outcomes study additional tables and figures

Definition for progression to diabetes Sample selection for outcomes analysis

