## Additional File 1. Implementation Study for "Effect of discussing personalized estimates of diabetes risk for people with prediabetes"

#### Supplementary Appendix

### Supplement 1. Patient Survey

**Pre- and Post- Dissemination Survey: PATIENTS**

1) What type of health care provider do you regularly see? Check one of the following:

   - O Primary care physician (a doctor that provides ongoing general care)
   - O Specialist (such as a heart doctor or diabetes doctor)
   - O Other, please explain: ________________________________________________

2) Has your doctor or nurse ever told you that you were at risk for getting diabetes? For example, they might have told you that you have high blood sugar, which may eventually lead to getting diabetes.

   Yes        No        Unsure

*If your answer to #2 was yes, please continue:*

3) If your doctor or nurse told you that you had a certain amount of risk for getting diabetes, what was the level of risk? Check one of the following:

   - O Low
   - O Medium
   - O High
   - O Percentage: ___________%
   - O Other, please explain: __________________________________________________
   - O Did not specify a level of risk

4) How would you prefer to get information about your risk of diabetes? Check all that apply:

   - ☐ Talking to your doctor
   - ☐ Talking to your nurse
   - ☐ Talking to another health care professional (such as a dietitian, pharmacist, or social worker)
   - ☐ Patient portal through your doctor's office
   - ☐ Getting information online
   - ☐ Reading materials
   - ☐ Cell phone app
   - ☐ Other, please explain: ___________________________________________________

5) Did your doctor or nurse mention or explain any tool(s) he or she used to figure out your level of risk for diabetes? For example, they might have used a computer program or an online calculator.

   Yes        No        Unsure        N/A (no risk discussion with provider)

6) On a scale from 1 to 5 (where 1 = "not at all confident" and 5 = "very confident"), how confident are you that you understand your risk of getting diabetes?

   Not at all confident   1        2        3        4        5   Very confident

7) If your doctor or nurse did use a tool, did it help you understand your risk?

- O Yes, it helped me to better understand my risk
- O No, it did not make any difference
- O Unsure
- O N/A (no calculator used)

8) Did your doctor or nurse recommend that you do anything to help avoid diabetes? For example, they might have suggested that you make lifestyle changes (such as going to the gym or starting a diet) or start medication.

Yes No Unsure N/A

9) Which action(s) did your doctor or nurse recommend? Check all that apply:

- ☐ Diet
- ☐ Exercise
- ☐ Medication (such as metformin)
- ☐ Diabetes Prevention Program (DPP)
- ☐ Other programs, please specify: _____________________________________________
- ☐ N/A (doctor/nurse did not recommend any actions)

10) Did you (or did you plan to) follow your doctor or nurse's advice and take any of the above actions? Check one of the following:

- O No, I do not intend to in the next 6 months
- O No, but I plan to in the next 6 months
- O Yes, I have taken action at least once in the past month
- O N/A (doctor/nurse did not recommend any actions)

11) If you did not take any action, why not? Check all that apply:

- ☐ I am planning to, but have not yet made any changes
- ☐ I did not believe I was at risk
- ☐ I believed I was at risk, but have not been ready or able to make changes
- ☐ I was unsure about my risk
- ☐ I could not afford to pay for medication or an organized lifestyle program
- ☐ I do not like to take medication
- ☐ I do not like to exercise, or it's too much work
- ☐ I am not worried about getting diabetes
- ☐ Other reason, please explain: _______________________________________________
- ☐ N/A (doctor/nurse did not recommend any actions)

12) On a scale from 1 to 5 (where 1 = "not at all satisfied" and 5 = "very satisfied"), how satisfied were you with the experience of talking to your provider about your risk of diabetes?

| 1 | 2 | 3 | 4 | 5 | N/A |
|---|---|---|---|---|---|
| Not at all satisfied | | | | Very Satisfied | |

### Supplement 2. Provider Surveys

**Pre-Dissemination Survey: PROVIDERS**

*For questions 1-4, please indicate how confident you are on a scale from 1 to 5, where 1 = "not at all confident" and 5 = "very confident."*

***How confident are you in your ability to:***

1) Estimate the average risk of diabetes progression for your patients with prediabetes?

   Not at all confident 1 2 3 4 5 Very confident

2) Identify clinical factors in your prediabetes patients that could increase or decrease their risk of diabetes progression?

   Not at all confident 1 2 3 4 5 Very confident

3) Directly communicate the degree of benefit a prediabetes patient should anticipate if they attend a DPP lifestyle modification program or take metformin?

   Not at all confident 1 2 3 4 5 Very confident

4) Tailor your recommendations for diabetes prevention to the general risk of an individual prediabetes patient?

   Not at all confident 1 2 3 4 5 Very confident

*For questions 5-7, please indicate how frequently you perform the indicated task.*

***How often do you:***

5) Inform prediabetes patients of their level of risk (low, medium, high, or numeric) of developing diabetes?

   None of the time Some of the time Most of the time All of the time

6) Recommend specific diabetes prevention interventions (lifestyle modification program or metformin) to prediabetes patients?

   None of the time Some of the time Most of the time All of the time

7) Refer patients to the Diabetes Prevention Program (DPP)?

   None of the time Some of the time Most of the time All of the time

*Please complete the rest of the survey (#8).*

8) How do you believe your patients would prefer to get information about their risk of diabetes?

- ○ Talking to their doctor
- ○ Talking to their nurse
- ○ Talking to another health care professional, e.g., dietitian, pharmacist, social worker
- ○ Patient portal through their doctor's office
- ○ Getting information online
- ○ Reading materials
- ○ Cell phone app
- ○ Other, please explain: ____________________________________________________

**Post-Dissemination Survey: PROVIDERS**

*For questions 1-5, please indicate how confident you are on a scale from 1 to 5, where 1 = "not at all confident" and 5 = "very confident."*

***How confident are you in your ability to:***

1) Estimate the average risk of diabetes progression for your patients with prediabetes?

   Not at all confident 1 2 3 4 5 Very confident

2) Identify clinical factors in your prediabetes patients that could increase or decrease their risk of diabetes progression?

   Not at all confident 1 2 3 4 5 Very confident

3) Directly communicate the degree of benefit a prediabetes patient should anticipate if they attend a DPP lifestyle modification program or take metformin?

   Not at all confident 1 2 3 4 5 Very confident

4) Tailor your recommendations for diabetes prevention to the general risk of an individual prediabetes patient?

   Not at all confident 1 2 3 4 5 Very confident

5) Using or tailoring the tool to convey information about a prediabetes patient's risk of diabetes progression?

   Not at all confident 1 2 3 4 5 Very confident

*For questions 6-8, please indicate how frequently you perform the indicated task.*

***How often do you:***

6) Inform your prediabetes patients of their level of risk (low, medium, high, or numeric) of developing diabetes?

   None of the time Some of the time Most of the time All of the time

7) Recommend specific diabetes prevention interventions (lifestyle modification or metformin) to your prediabetes patients?

   None of the time Some of the time Most of the time All of the time

8) Refer patients to the Diabetes Prevention Program (DPP)?

   None of the time Some of the time Most of the time All of the time

*Please complete the rest of the survey (#9-20).*

9) Did you receive information or training on how to use the Tufts DPP tool?

   Yes No Unsure

10) Do you feel competent in the use of the Tufts DPP tool?

   Yes No Unsure

11) How often could you use the tool to screen patients at risk for diabetes?

   None of the time Some of the time Most of the time All of the time

12) How often have you used the tool to screen patients at risk for diabetes?

   None of the time Some of the time Most of the time All of the time

13) To what degree did using the tool heighten your awareness of the heterogeneity of risk of diabetes among prediabetes patients?

   Not at all A little Some A great deal

14) To what degree did using the tool heighten your awareness of heterogeneity of individual benefit from specific therapies among prediabetes patients?

   Not at all A little Some A great deal

15) Has using this tool affected the quality of your encounters with patients with prediabetes?

   Yes, improved quality No effect Yes, decreased quality

16) In your opinion, has using this tool influenced patients' decisions to adhere to medication or enroll in the DPP?

   Yes, positive impact No effect Yes, negative impact

17) Has using this tool impacted your work protocols and flow when seeing patients with prediabetes?

   Yes, positive impact No effect Yes, negative impact

18) To what degree has this tool influenced how likely you are to recommend a diabetes prevention intervention (DPP lifestyle intervention or metformin) for prediabetes patients?

   Not at all A little Some A great deal

19) To what degree has this tool influenced your decisions regarding which prediabetes patients you will personally maintain and refer to a diabetes prevention intervention (DPP lifestyle intervention or metformin)?

Not at all A little Some A great deal

20) How do you believe your patients would prefer to get information about their risk of diabetes?
- Talking to their doctor
- Talking to their nurse
- Talking to another health care professional, e.g., dietitian, pharmacist, social worker
- Patient portal through their doctor's office
- Getting information online
- Reading materials
- Cell phone app
- Other, please explain: ______________________________________________

### Supplement 3. Premier Patient Survey Results

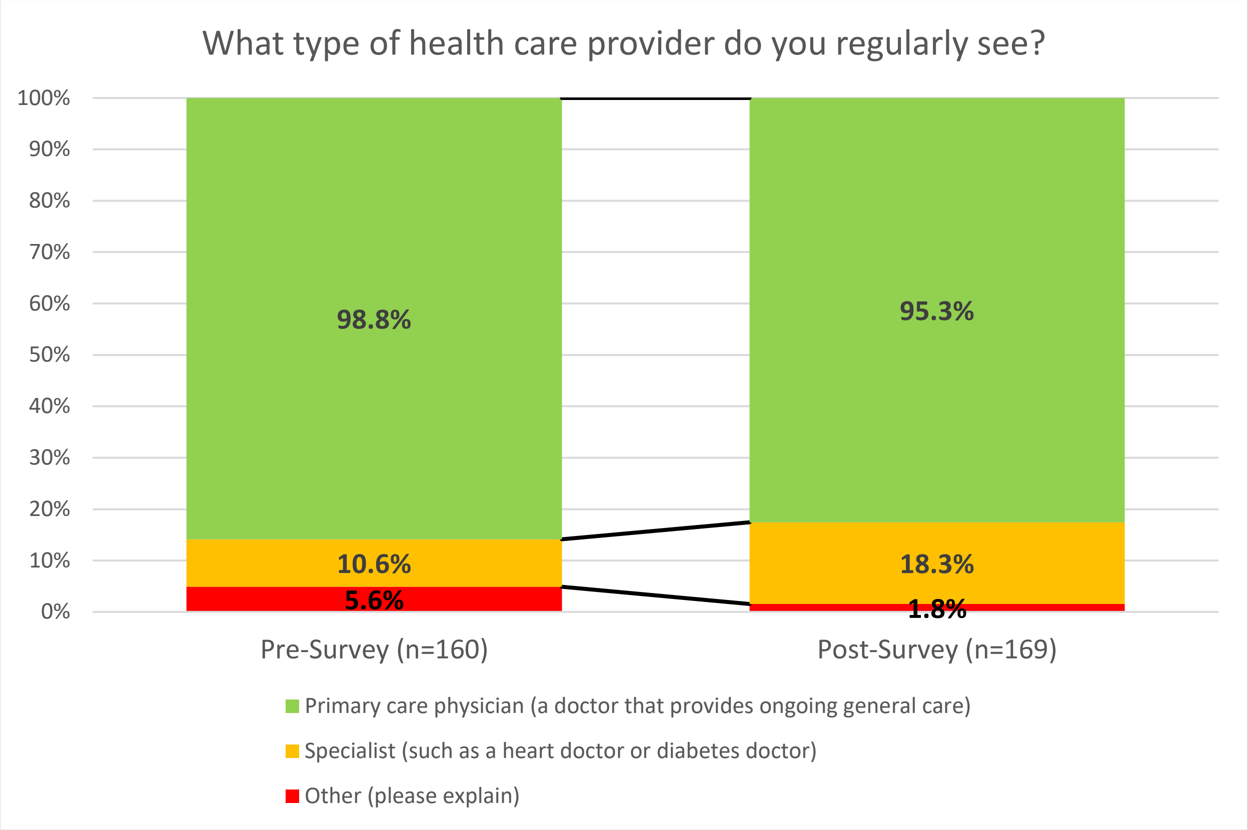

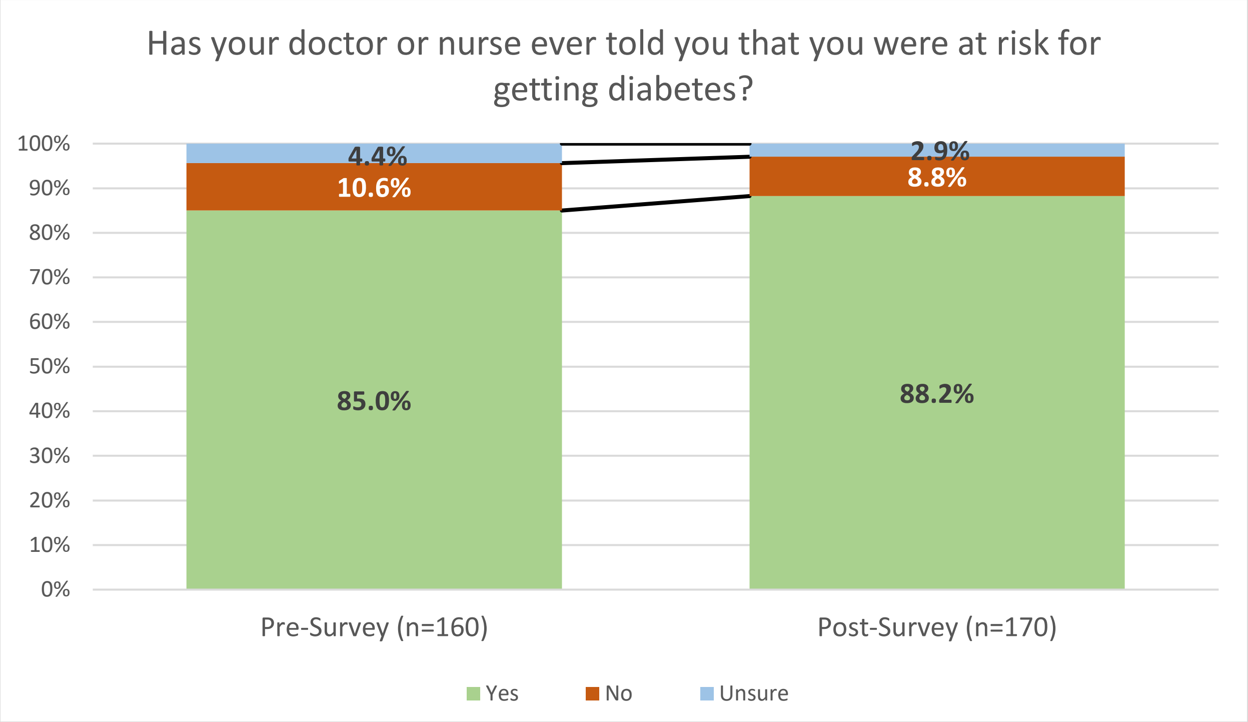

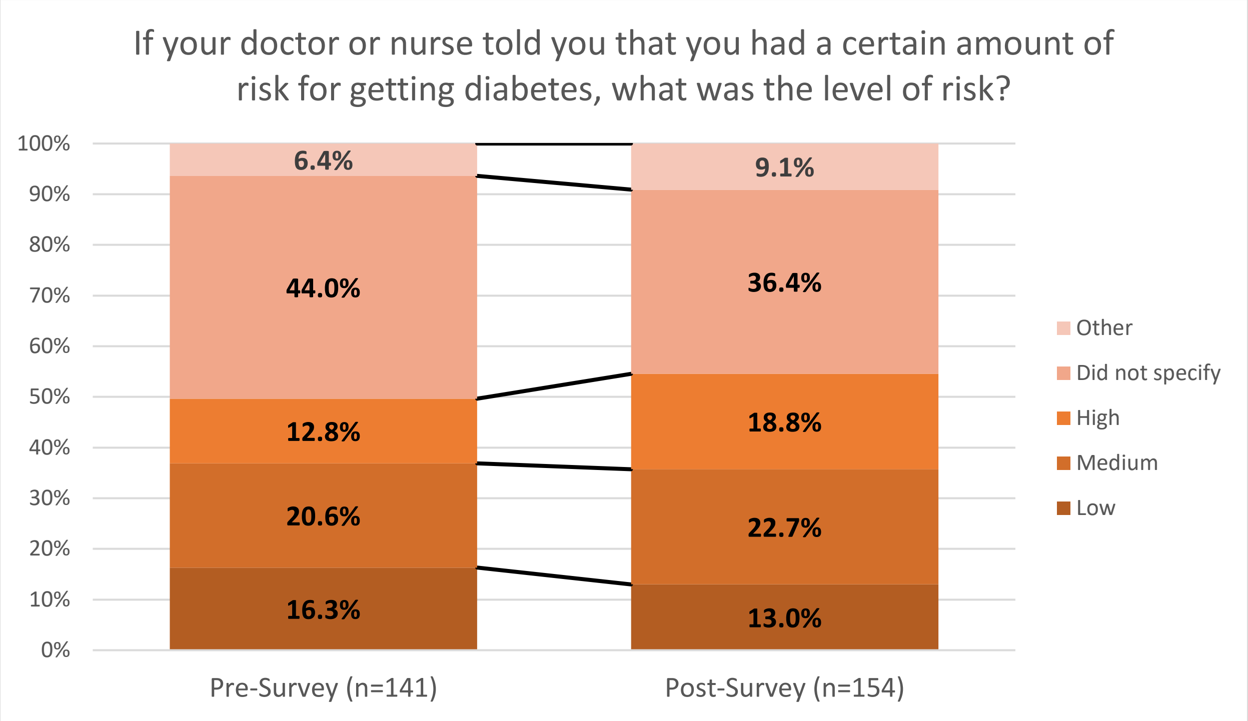
If your doctor or nurse told you that you had a certain amount of risk for getting diabetes, what was the level of risk?
100%
90%
80%
70%
60%
50%
40%
30%
20%
10%
0%
6.4%
44.0%
12.8%
20.6%
16.3%
9.1%
36.4%
18.8%
22.7%
13.0%
Other
Did not specify
High
Medium
Low
Pre-Survey (n=141)
Post-Survey (n=154)

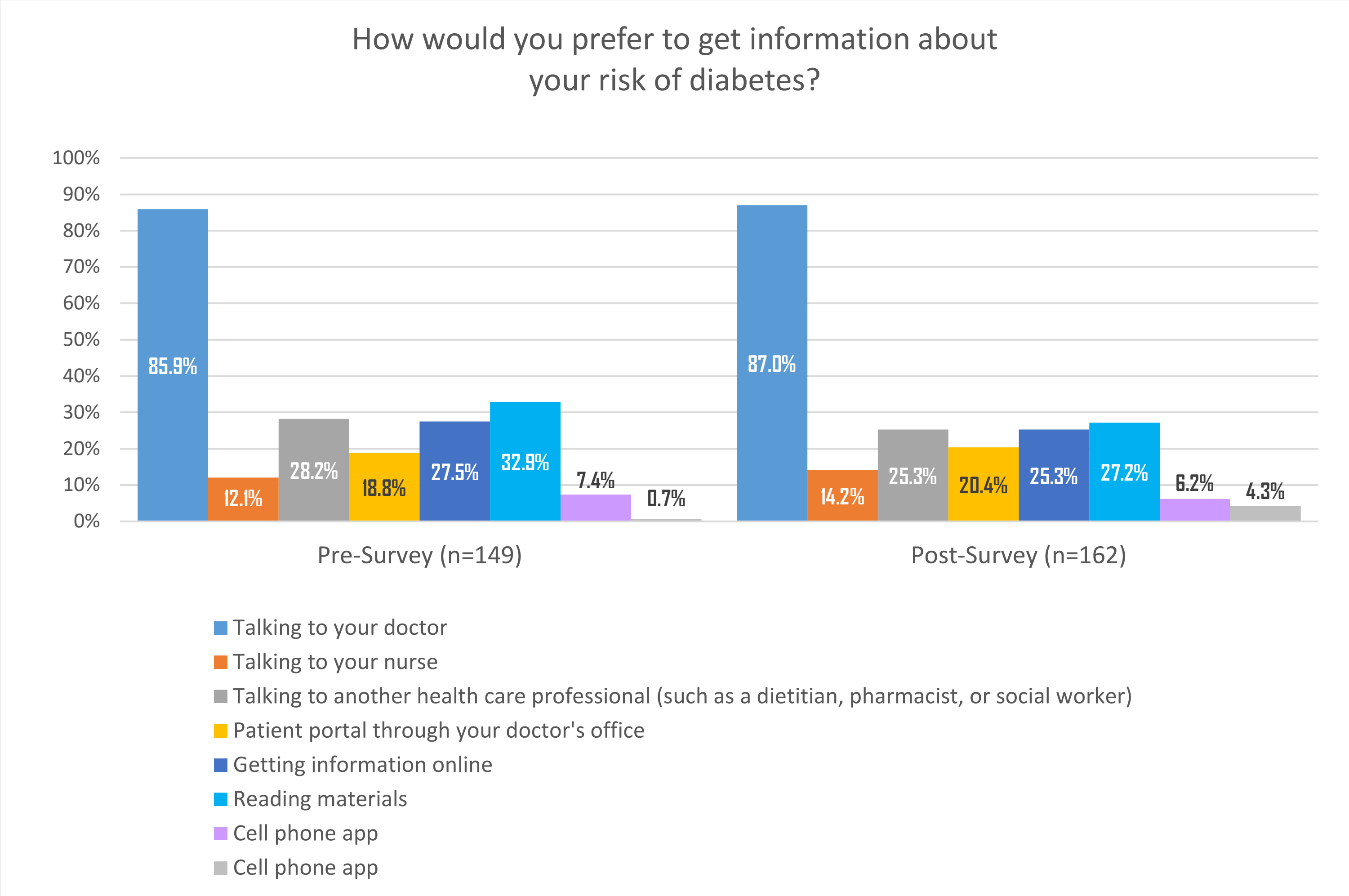
How would you prefer to get information about your risk of diabetes?
100%
90%
80%
70%
60%
50%
40%
30%
20%
10%
0%
85.9%
12.1%
28.2%
18.8%
27.5%
32.9%
7.4%
0.7%
87.0%
14.2%
25.3%
20.4%
25.3%
27.2%
6.2%
4.3%
Pre-Survey (n=149)
Post-Survey (n=162)
Talking to your doctor
Talking to your nurse
Talking to another health care professional (such as a dietitian, pharmacist, or social worker)
Patient portal through your doctor's office
Getting information online
Reading materials
Cell phone app
Cell phone app

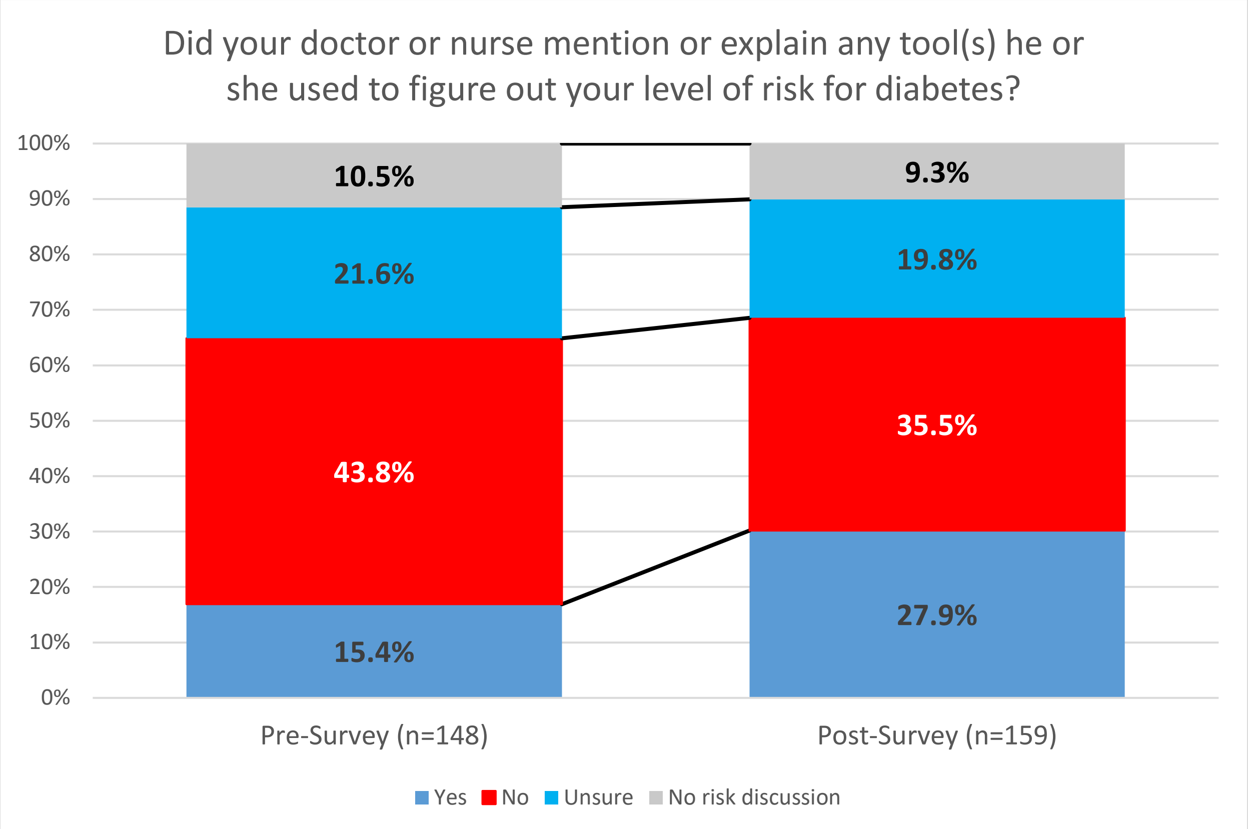
Did your doctor or nurse mention or explain any tool(s) he or she used to figure out your level of risk for diabetes?
100%
90%
80%
70%
60%
50%
40%
30%
20%
10%
0%
10.5%
21.6%
43.8%
15.4%
9.3%
19.8%
35.5%
27.9%
Pre-Survey (n=148)
Post-Survey (n=159)
Yes
No
Unsure
No risk discussion

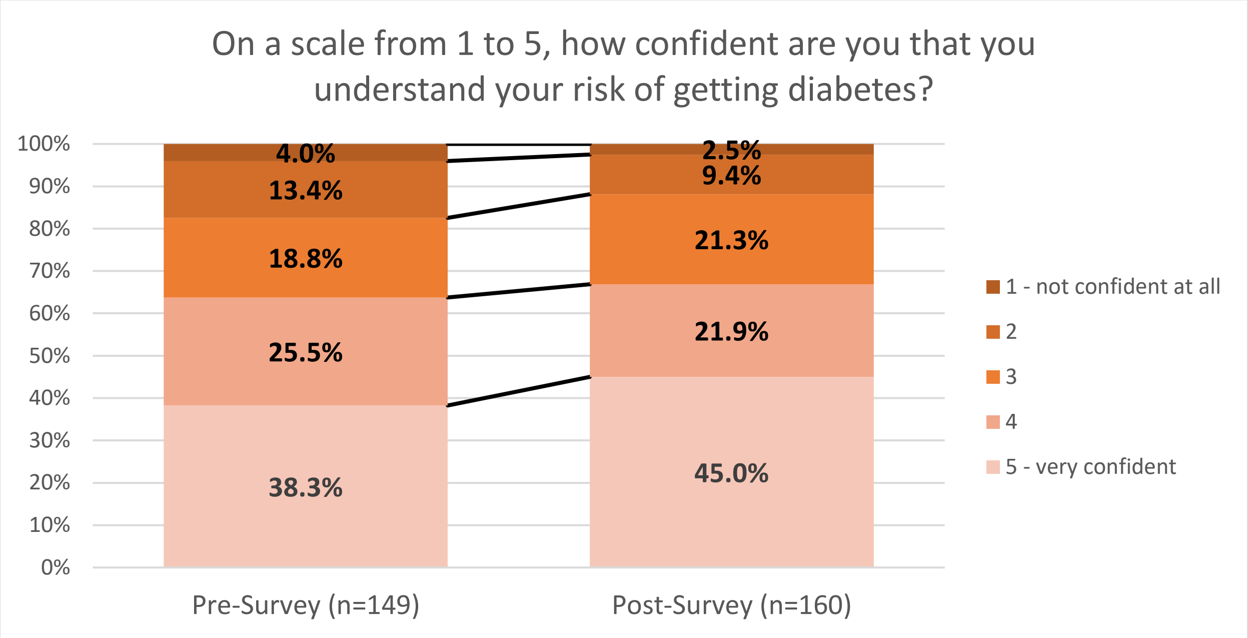
On a scale from 1 to 5, how confident are you that you understand your risk of getting diabetes?
100%
90%
80%
70%
60%
50%
40%
30%
20%
10%
0%
4.0%
13.4%
18.8%
25.5%
38.3%
2.5%
9.4%
21.3%
21.9%
45.0%
Pre-Survey (n=149)
Post-Survey (n=160)
1 - not confident at all
2
3
4
5 - very confident

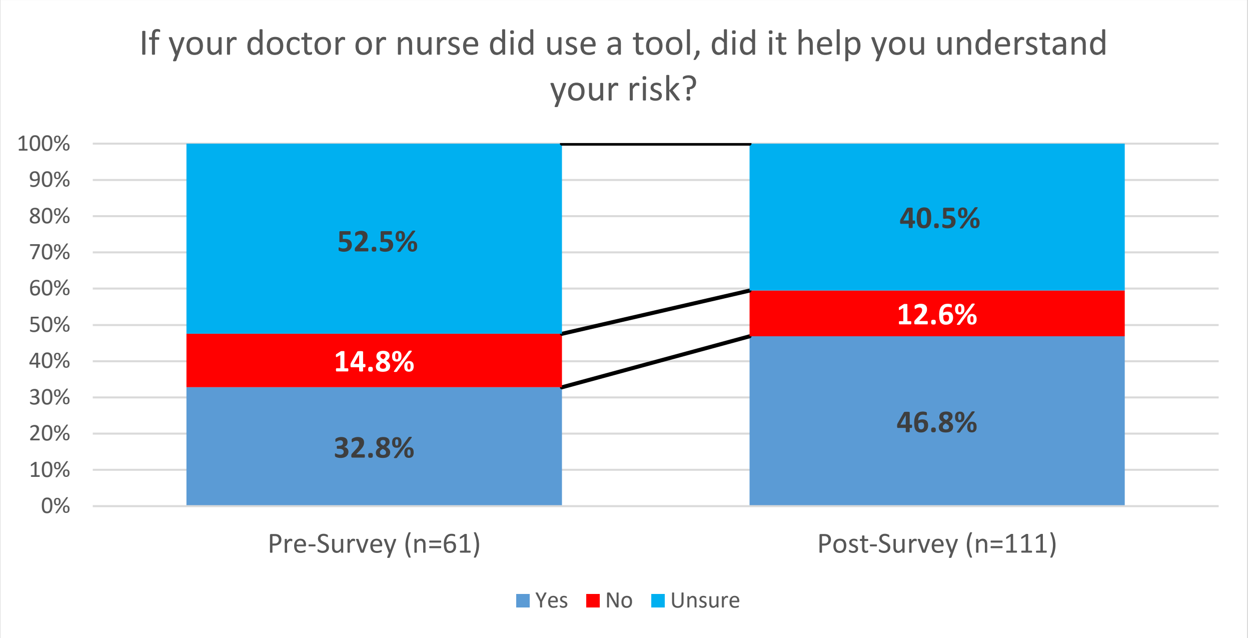
If your doctor or nurse did use a tool, did it help you understand your risk?
100%
90%
80%
70%
60%
50%
40%
30%
20%
10%
0%
52.5%
14.8%
32.8%
40.5%
12.6%
46.8%
Pre-Survey (n=61)
Post-Survey (n=111)
Yes
No
Unsure

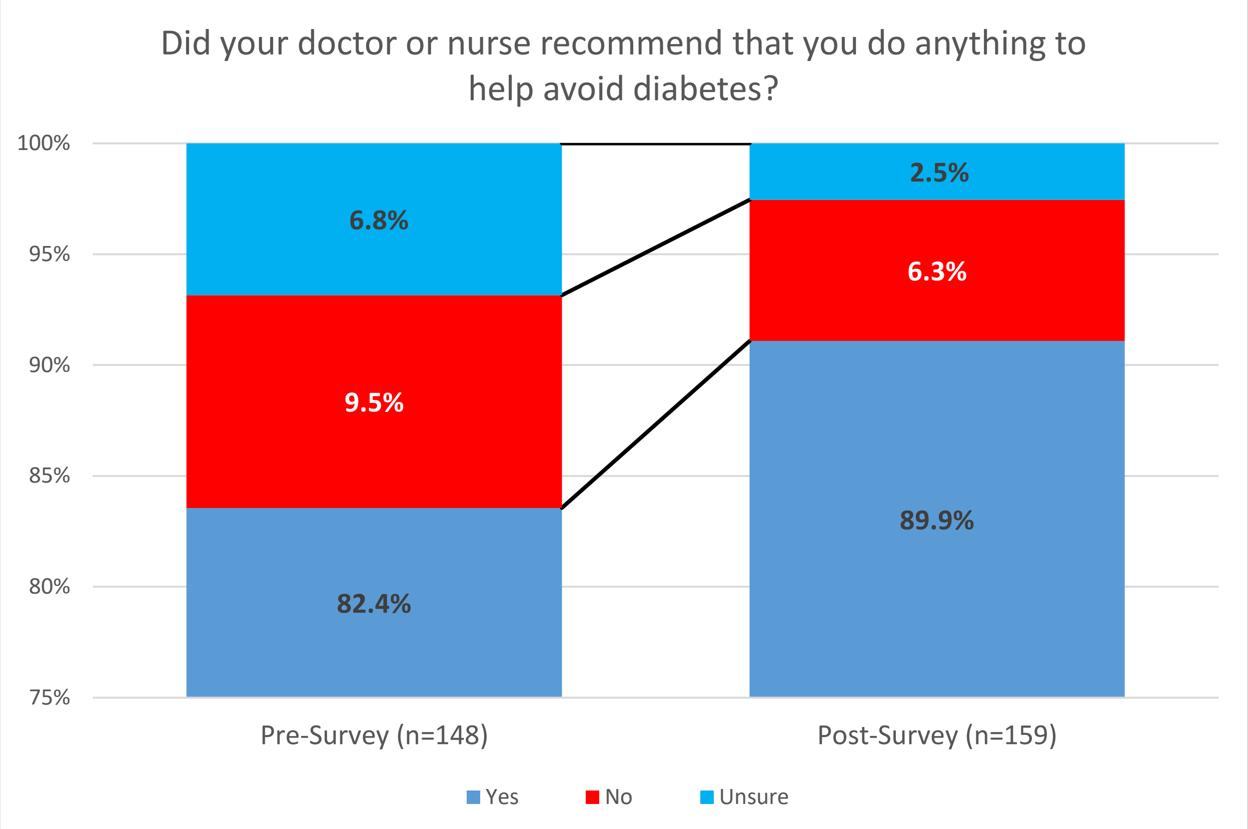
Did your doctor or nurse recommend that you do anything to help avoid diabetes?
100%
95%
90%
85%
80%
75%
6.8%
9.5%
82.4%
2.5%
6.3%
89.9%
Pre-Survey (n=148)
Post-Survey (n=159)
Yes
No
Unsure

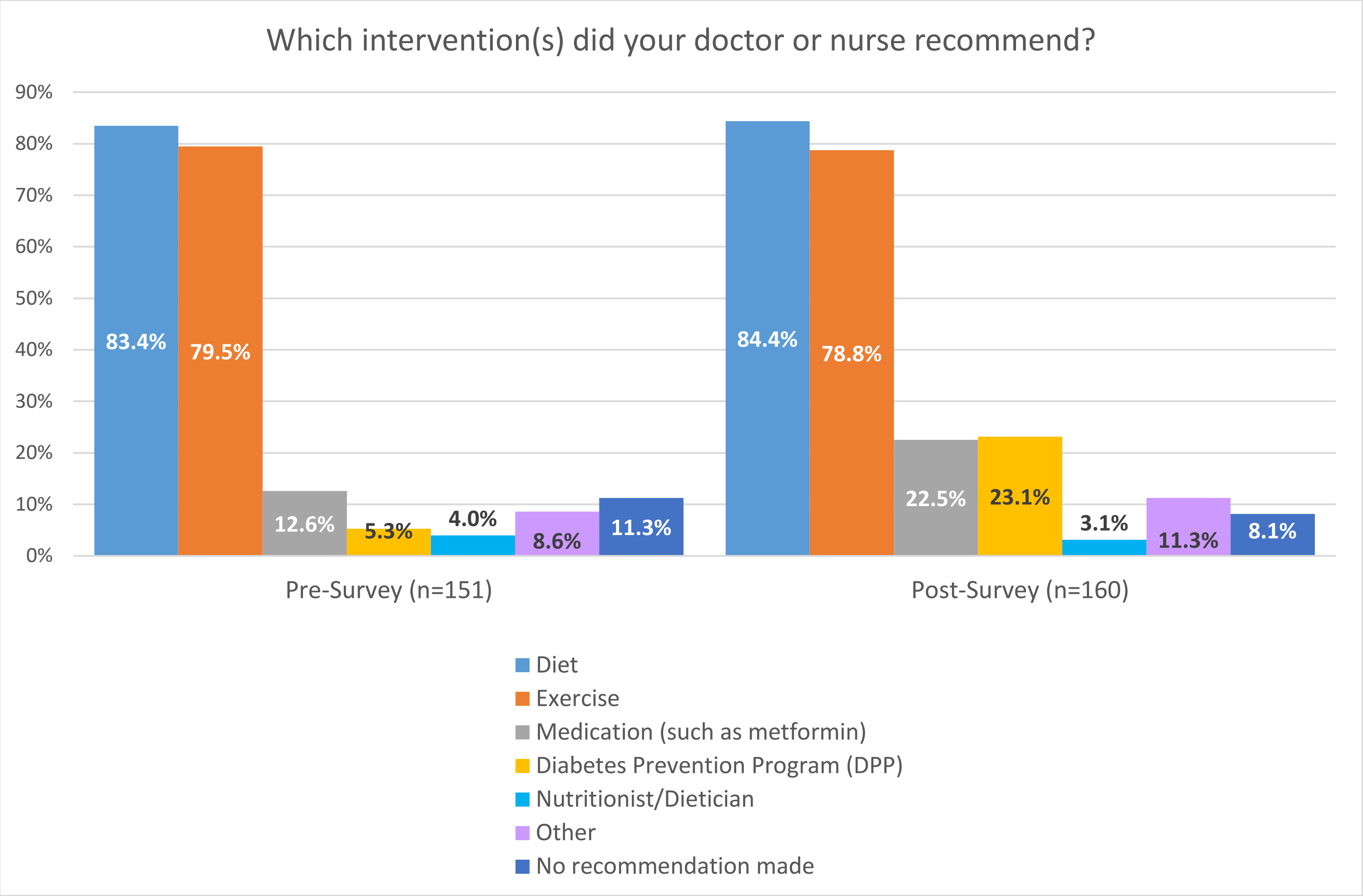
Which intervention(s) did your doctor or nurse recommend?
90%
80%
70%
60%
50%
40%
30%
20%
10%
0%
83.4%
79.5%
12.6%
5.3%
4.0%
8.6%
11.3%
84.4%
78.8%
22.5%
23.1%
3.1%
11.3%
8.1%
Pre-Survey (n=151)
Post-Survey (n=160)
Diet
Exercise
Medication (such as metformin)
Diabetes Prevention Program (DPP)
Nutritionist/Dietician
Other
No recommendation made

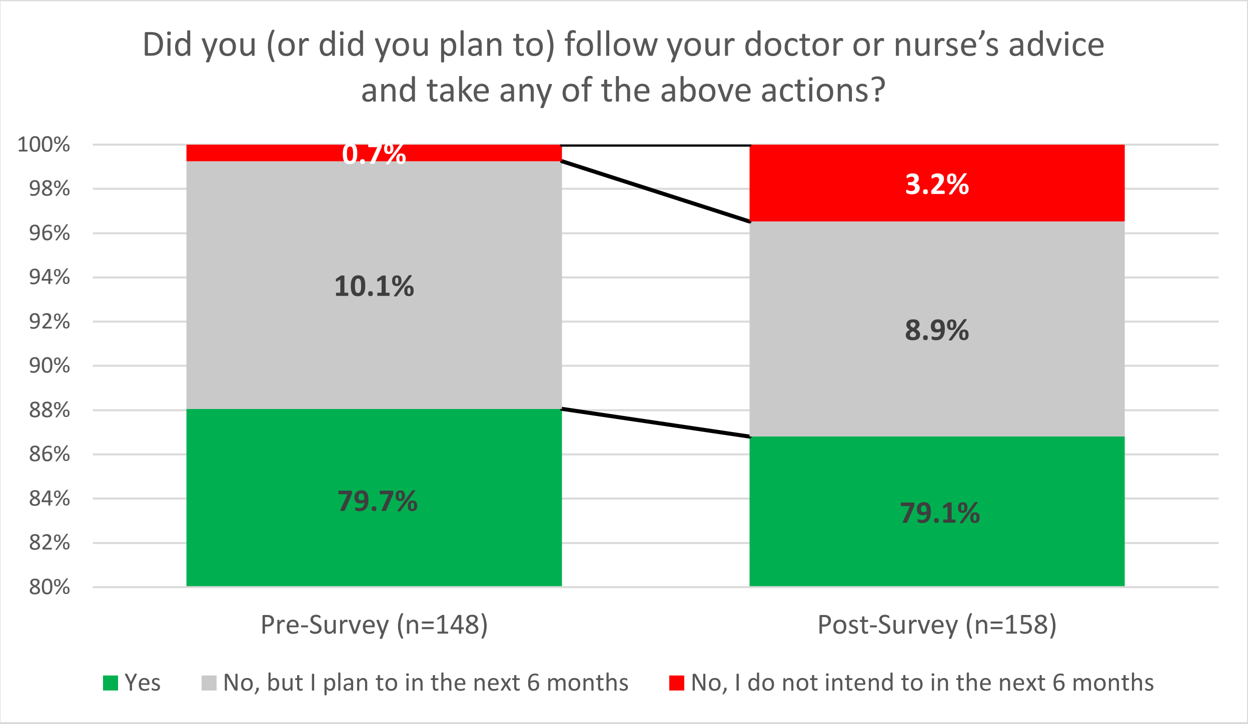
Did you (or did you plan to) follow your doctor or nurse's advice and take any of the above actions?
100%
98%
96%
94%
92%
90%
88%
86%
84%
82%
80%
0.7%
10.1%
79.7%
3.2%
8.9%
79.1%
Pre-Survey (n=148)
Post-Survey (n=158)
Yes
No, but I plan to in the next 6 months
No, I do not intend to in the next 6 months

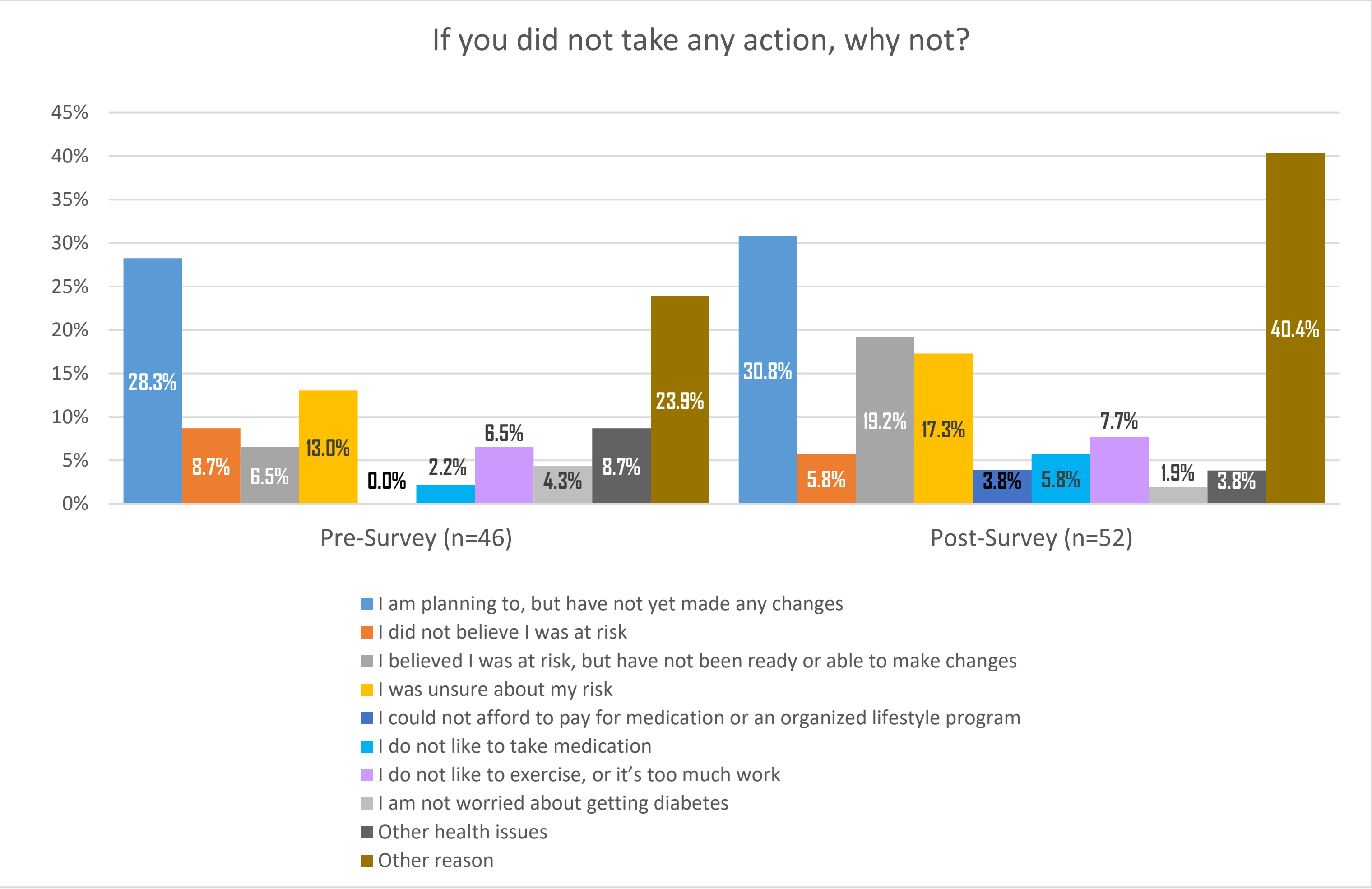
If you did not take any action, why not?
45%
40%
35%
30%
25%
20%
15%
10%
5%
0%
28.3%
8.7%
6.5%
13.0%
0.0%
2.2%
6.5%
4.3%
8.7%
23.9%
30.8%
5.8%
19.2%
17.3%
3.8%
5.8%
7.7%
1.9%
3.8%
40.4%
Pre-Survey (n=46)
Post-Survey (n=52)
I am planning to, but have not yet made any changes
I did not believe I was at risk
I believed I was at risk, but have not been ready or able to make changes
I was unsure about my risk
I could not afford to pay for medication or an organized lifestyle program
I do not like to take medication
I do not like to exercise, or it's too much work
I am not worried about getting diabetes
Other health issues
Other reason

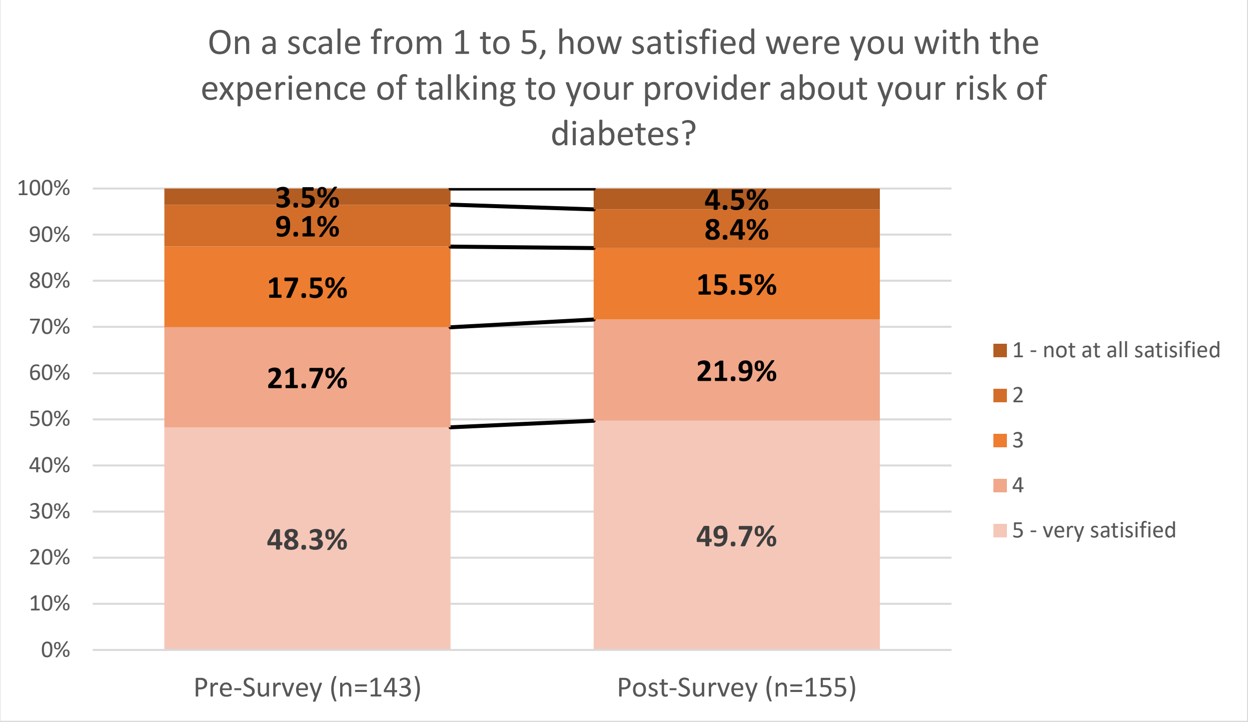
On a scale from 1 to 5, how satisfied were you with the experience of talking to your provider about your risk of diabetes?
100%
90%
80%
70%
60%
50%
40%
30%
20%
10%
0%
3.5%
9.1%
17.5%
21.7%
48.3%
4.5%
8.4%
15.5%
21.9%
49.7%
Pre-Survey (n=143)
Post-Survey (n=155)
1 - not at all satisified
2
3
4
5 - very satisified

#### Supplement 4. Premier Provider Survey Results

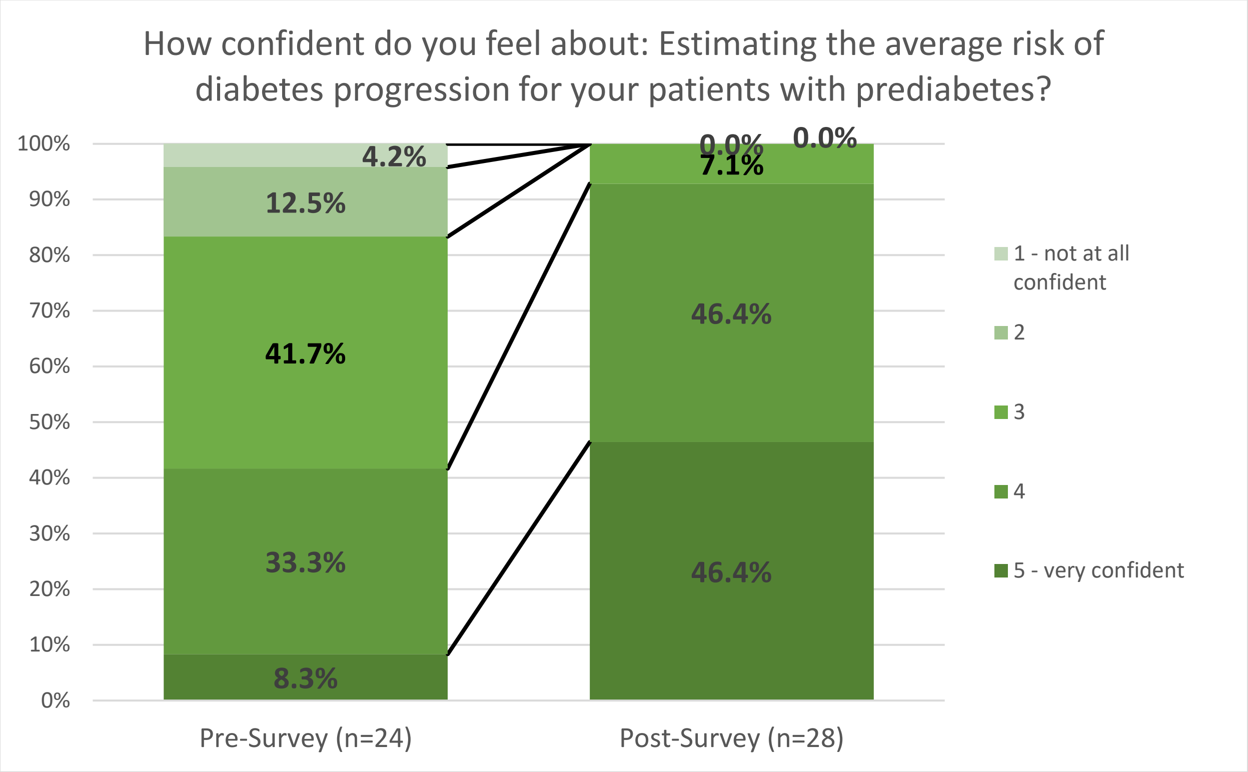

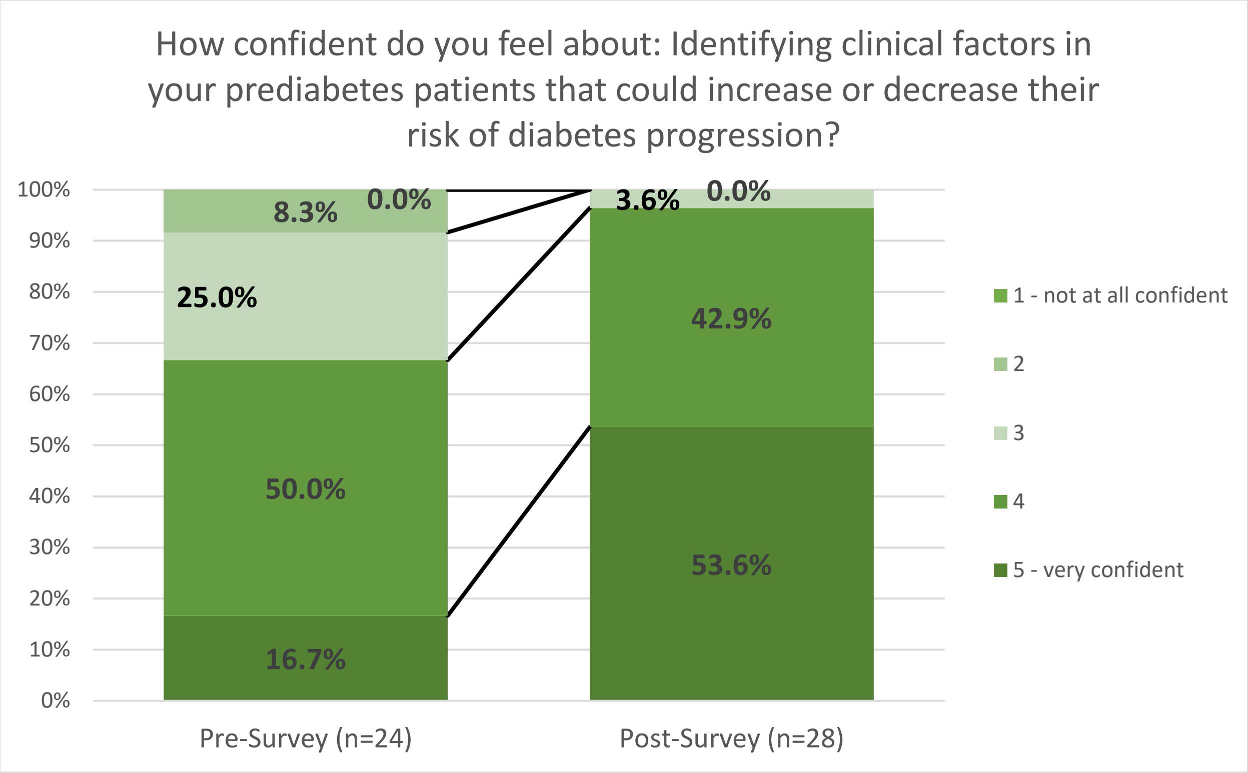

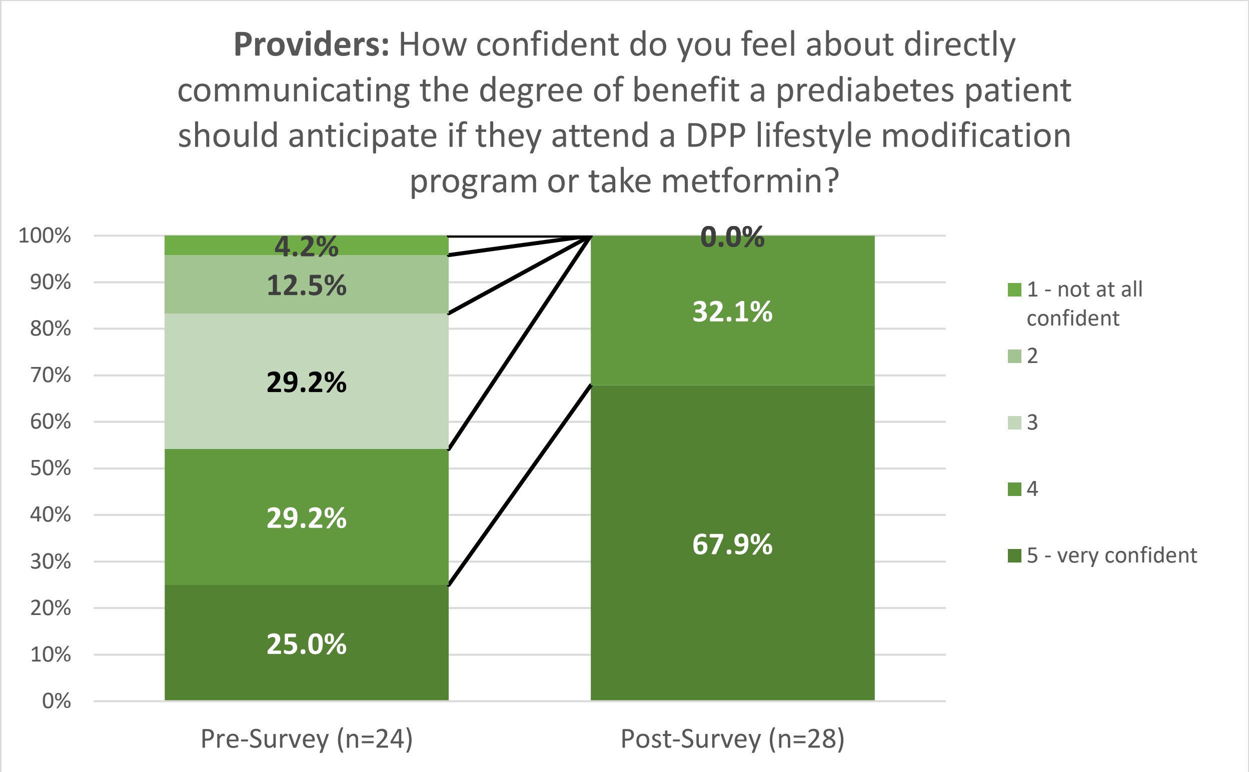
Providers: How confident do you feel about directly communicating the degree of benefit a prediabetes patient should anticipate if they attend a DPP lifestyle modification program or take metformin?
100%
90%
80%
70%
60%
50%
40%
30%
20%
10%
0%
4.2%
12.5%
29.2%
29.2%
25.0%
0.0%
32.1%
67.9%
Pre-Survey (n=24)
Post-Survey (n=28)
1 - not at all confident
2
3
4
5 - very confident

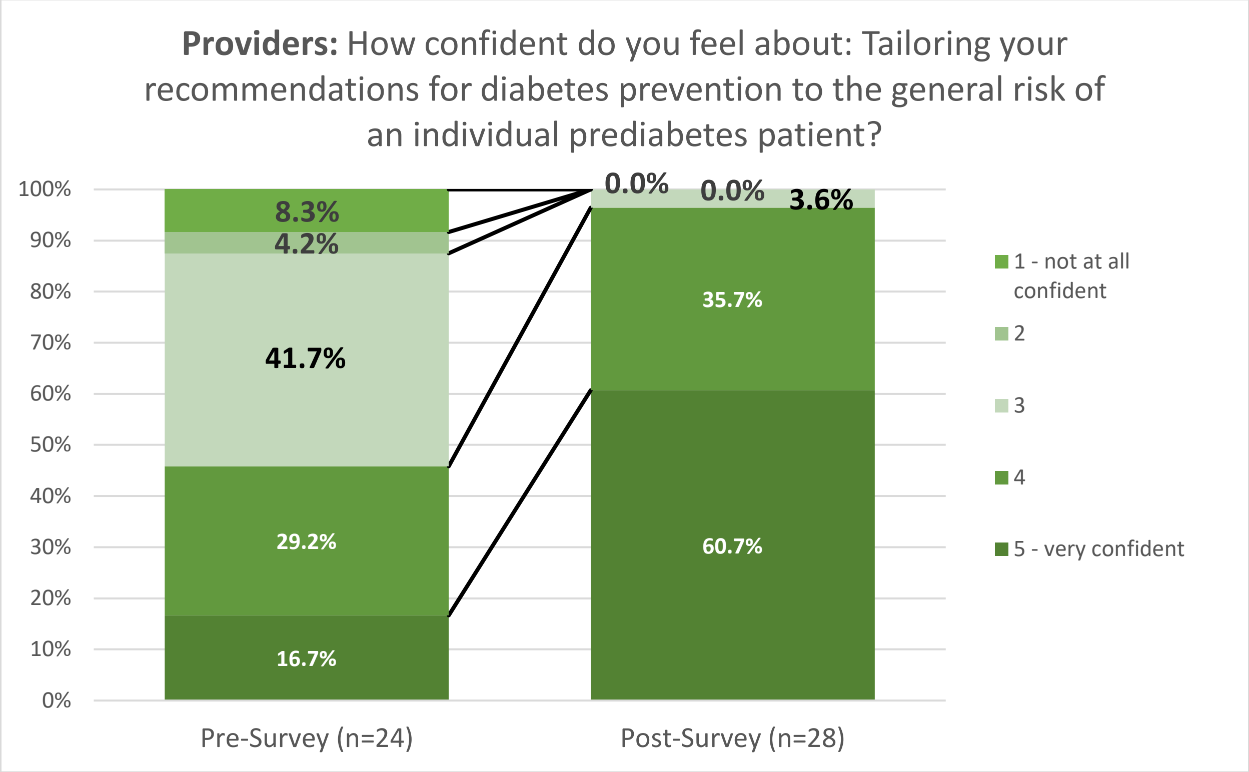
Providers: How confident do you feel about: Tailoring your recommendations for diabetes prevention to the general risk of an individual prediabetes patient?
100%
90%
80%
70%
60%
50%
40%
30%
20%
10%
0%
8.3%
4.2%
41.7%
29.2%
16.7%
0.0%
0.0%
3.6%
35.7%
60.7%
Pre-Survey (n=24)
Post-Survey (n=28)
1 - not at all confident
2
3
4
5 - very confident

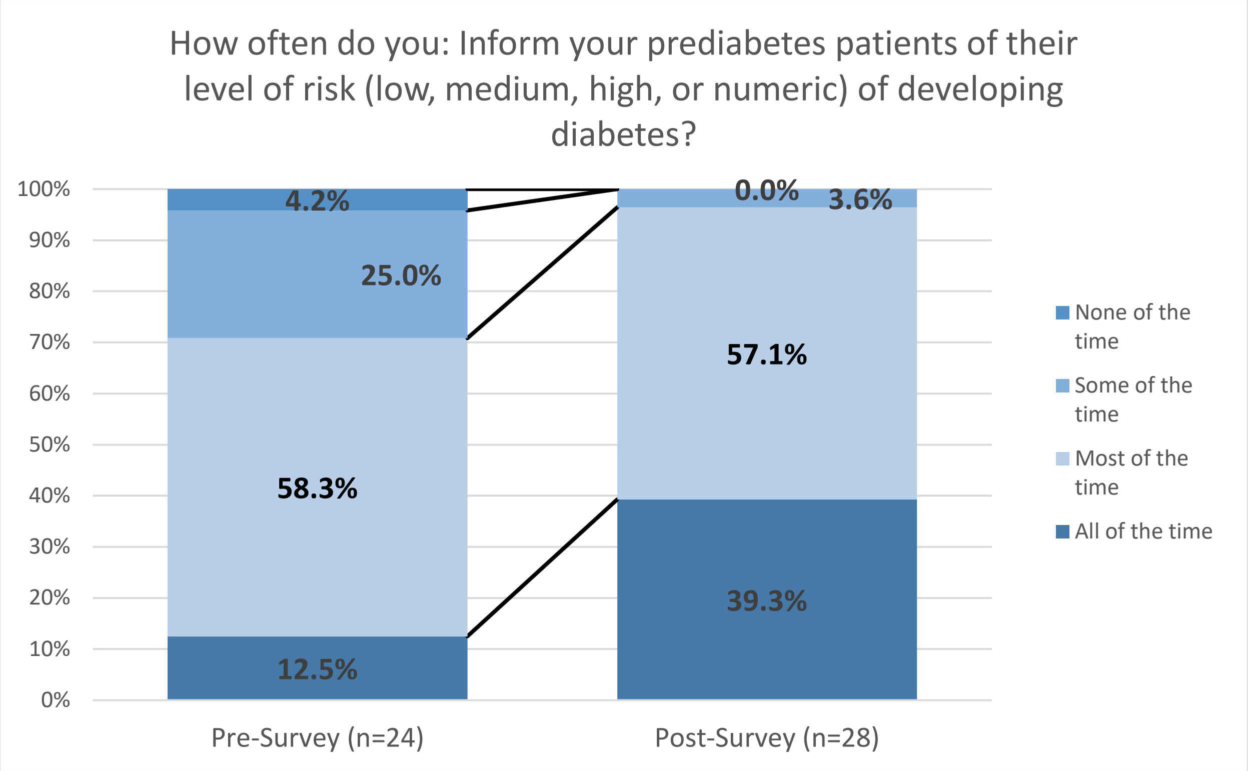
How often do you: Inform your prediabetes patients of their level of risk (low, medium, high, or numeric) of developing diabetes?
100%
90%
80%
70%
60%
50%
40%
30%
20%
10%
0%
4.2%
25.0%
58.3%
12.5%
0.0%
3.6%
57.1%
39.3%
Pre-Survey (n=24)
Post-Survey (n=28)
None of the time
Some of the time
Most of the time
All of the time

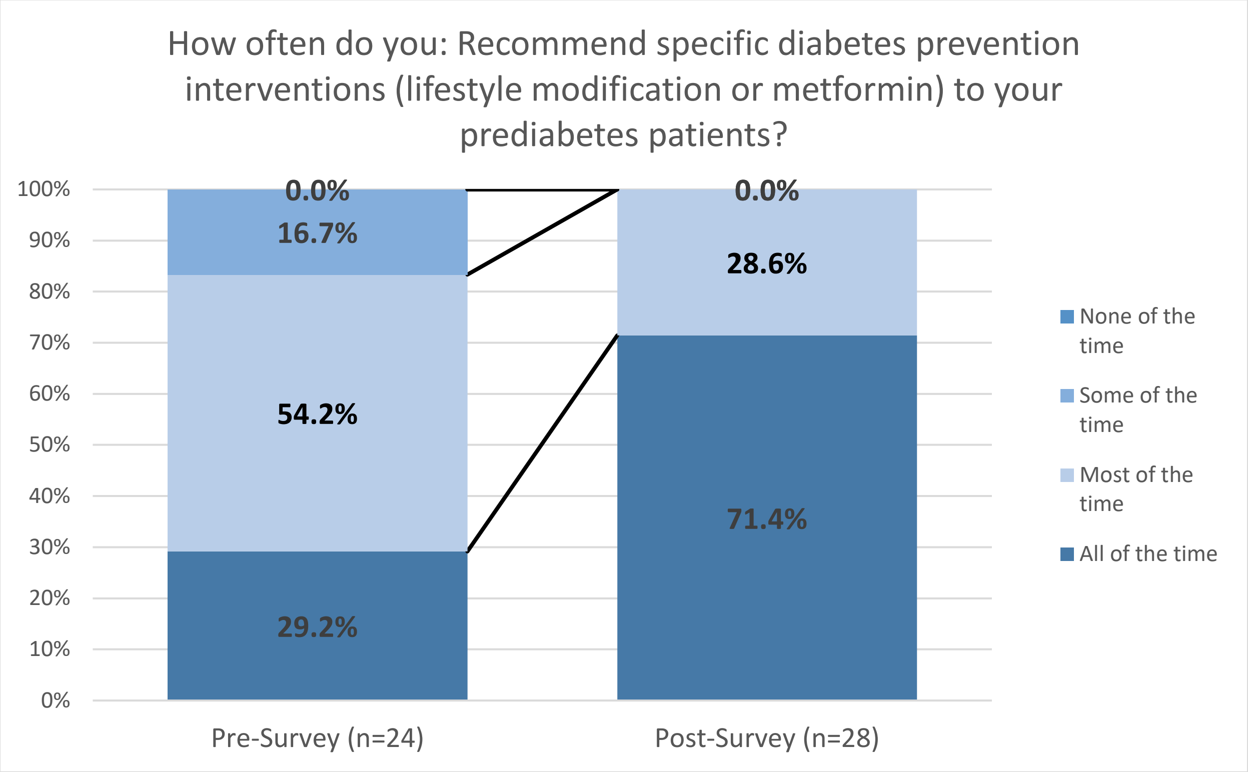
How often do you: Recommend specific diabetes prevention interventions (lifestyle modification or metformin) to your prediabetes patients?
100%
90%
80%
70%
60%
50%
40%
30%
20%
10%
0%
0.0%
16.7%
54.2%
29.2%
0.0%
28.6%
71.4%
Pre-Survey (n=24)
Post-Survey (n=28)
None of the time
Some of the time
Most of the time
All of the time

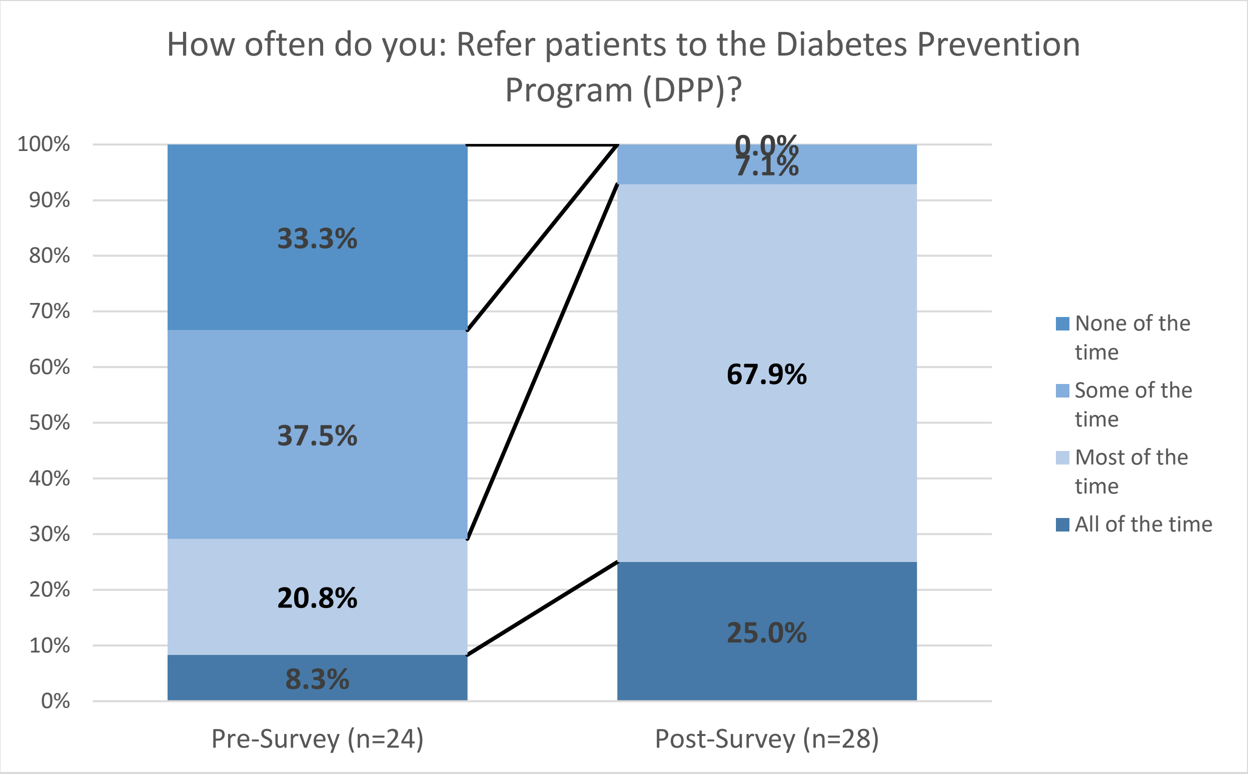
How often do you: Refer patients to the Diabetes Prevention Program (DPP)?
100%
90%
80%
70%
60%
50%
40%
30%
20%
10%
0%
33.3%
37.5%
20.8%
8.3%
0.0%
7.1%
67.9%
25.0%
Pre-Survey (n=24)
Post-Survey (n=28)
None of the time
Some of the time
Most of the time
All of the time

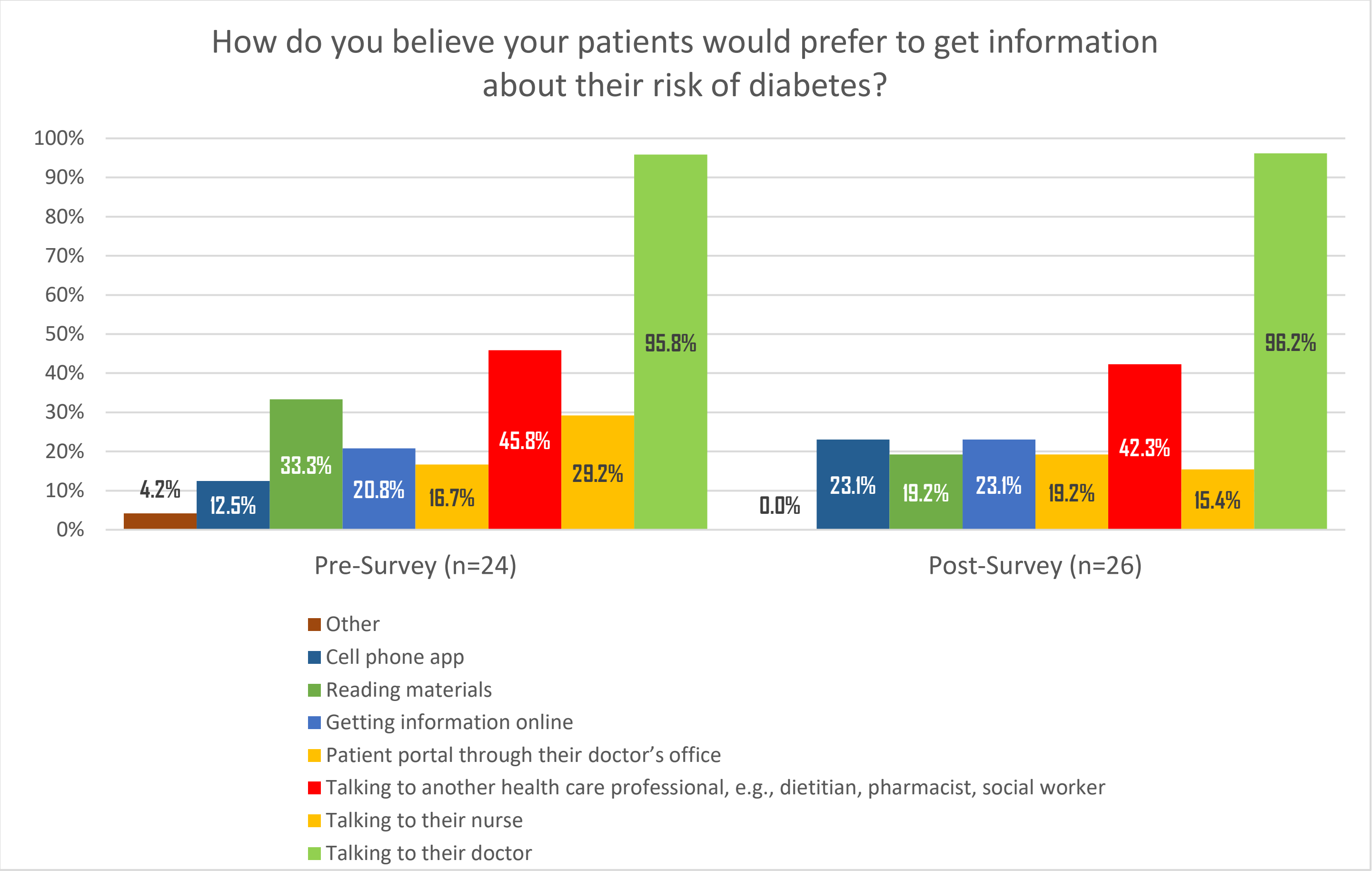
How do you believe your patients would prefer to get information about their risk of diabetes?
100%
90%
80%
70%
60%
50%
40%
30%
20%
10%
0%
4.2%
12.5%
33.3%
20.8%
16.7%
45.8%
29.2%
95.8%
0.0%
23.1%
19.2%
23.1%
19.2%
42.3%
15.4%
96.2%
Pre-Survey (n=24)
Post-Survey (n=26)
Other
Cell phone app
Reading materials
Getting information online
Patient portal through their doctor's office
Talking to another health care professional, e.g., dietitian, pharmacist, social worker
Talking to their nurse
Talking to their doctor

**<u>Post Survey only</u>**

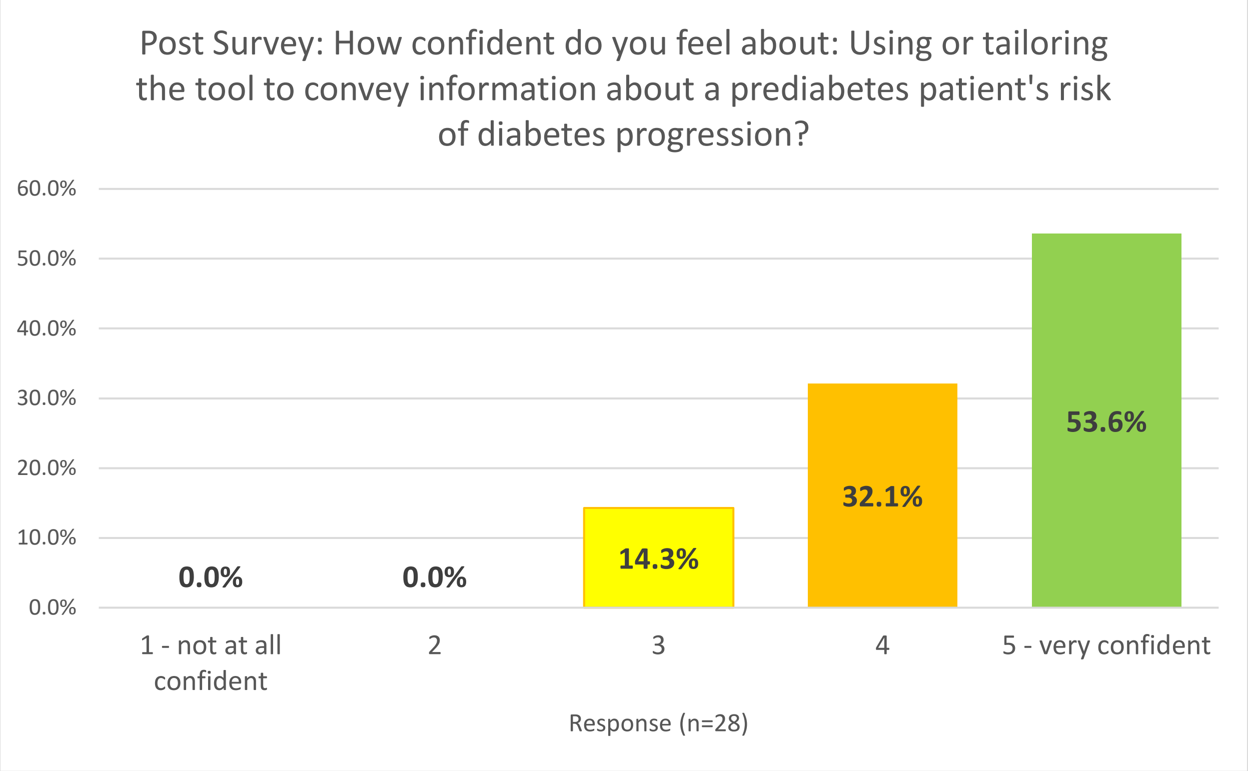

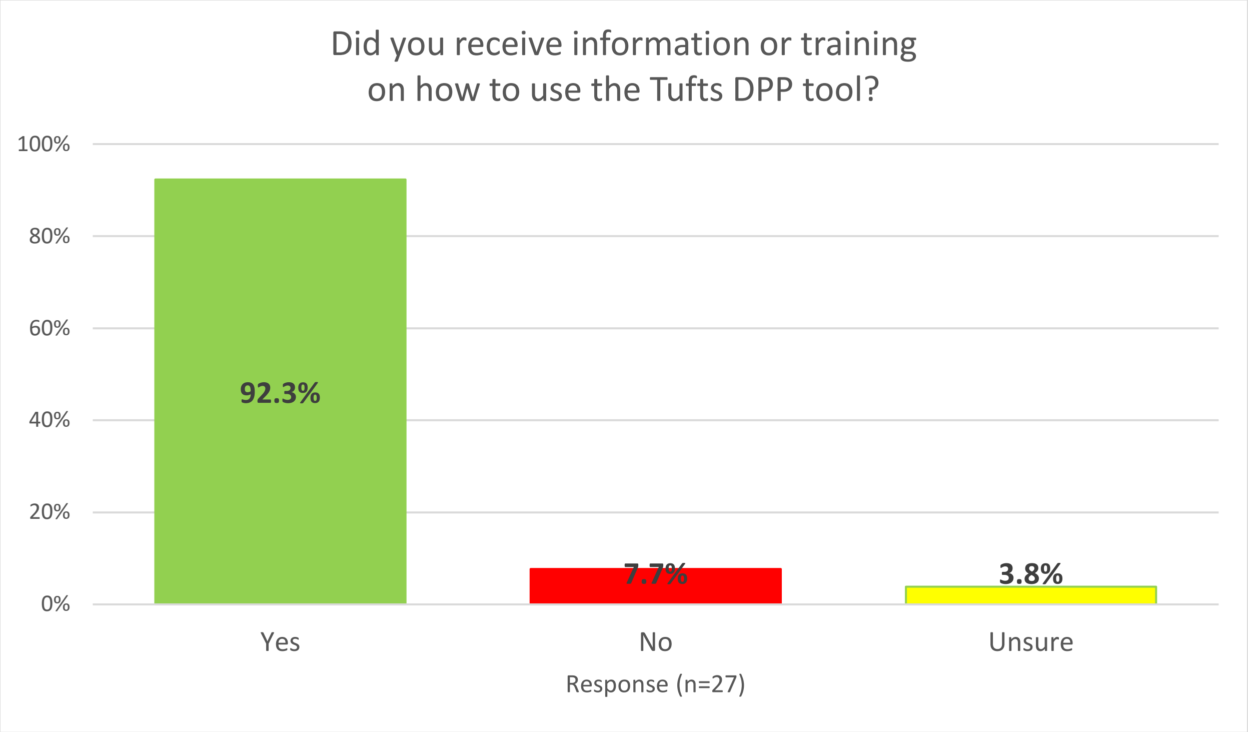

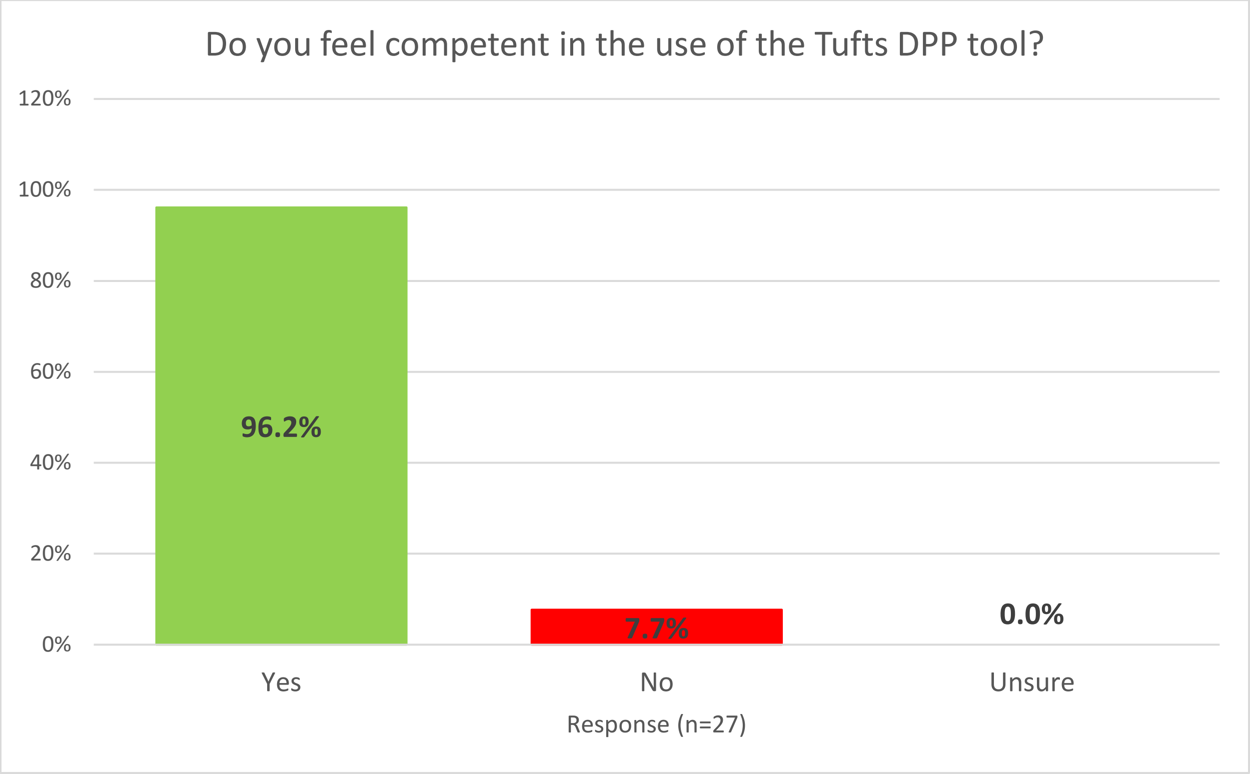
Do you feel competent in the use of the Tufts DPP tool?
120%
100%
80%
60%
40%
20%
0%
96.2%
7.7%
0.0%
Yes
No
Unsure
Response (n=27)

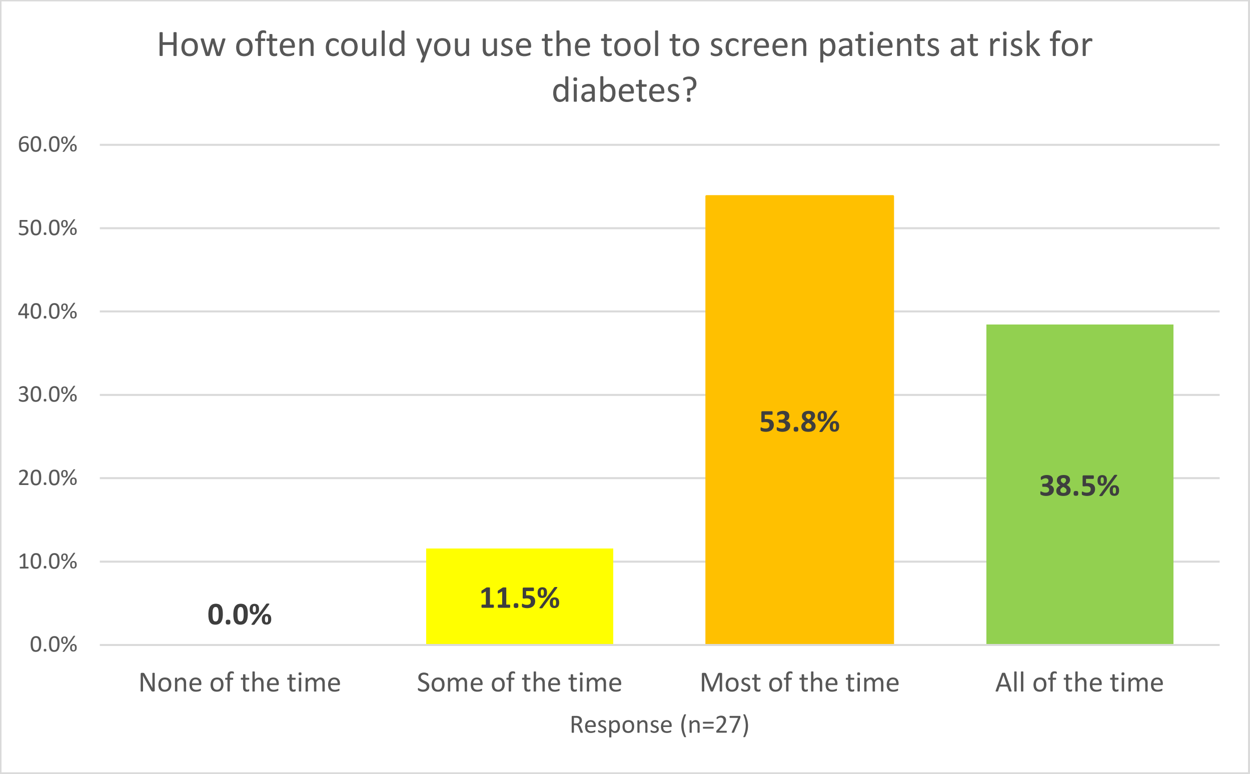
How often could you use the tool to screen patients at risk for diabetes?
60.0%
50.0%
40.0%
30.0%
20.0%
10.0%
0.0%
0.0%
11.5%
53.8%
38.5%
None of the time
Some of the time
Most of the time
All of the time
Response (n=27)

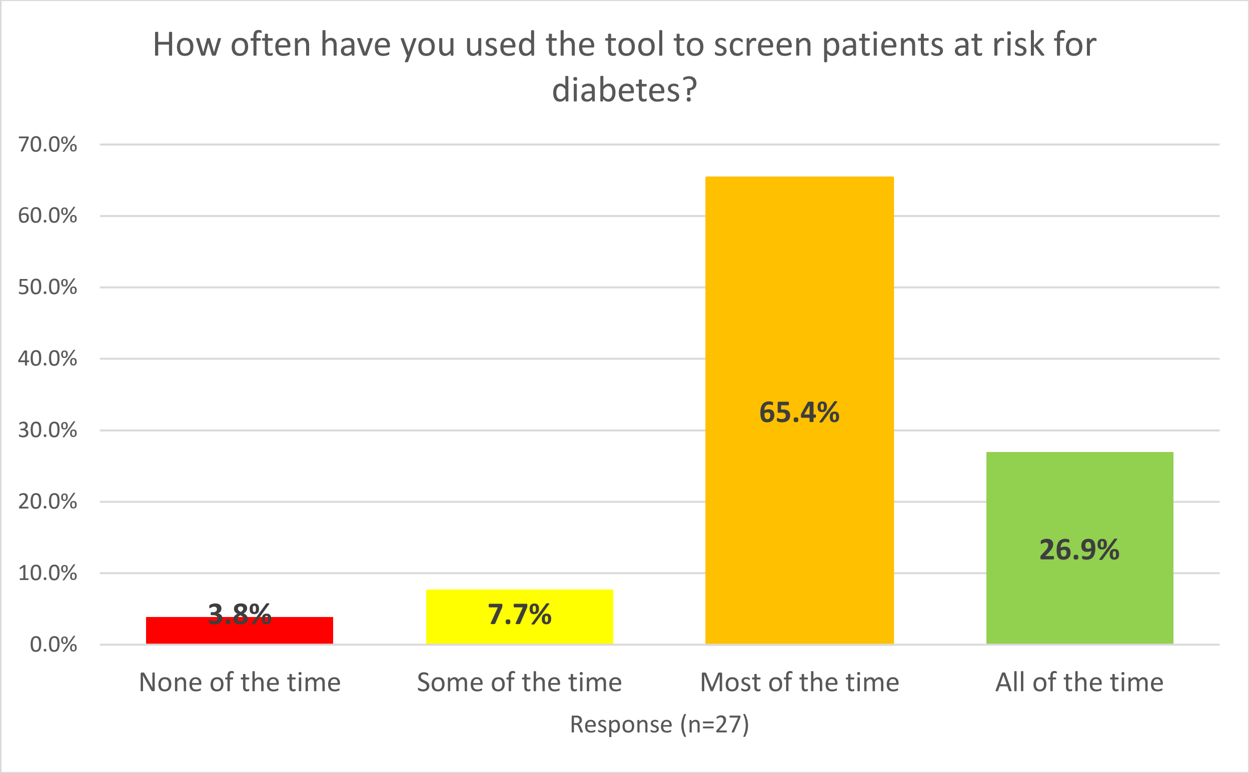
How often have you used the tool to screen patients at risk for diabetes?
70.0%
60.0%
50.0%
40.0%
30.0%
20.0%
10.0%
0.0%
3.8%
7.7%
65.4%
26.9%
None of the time
Some of the time
Most of the time
All of the time
Response (n=27)

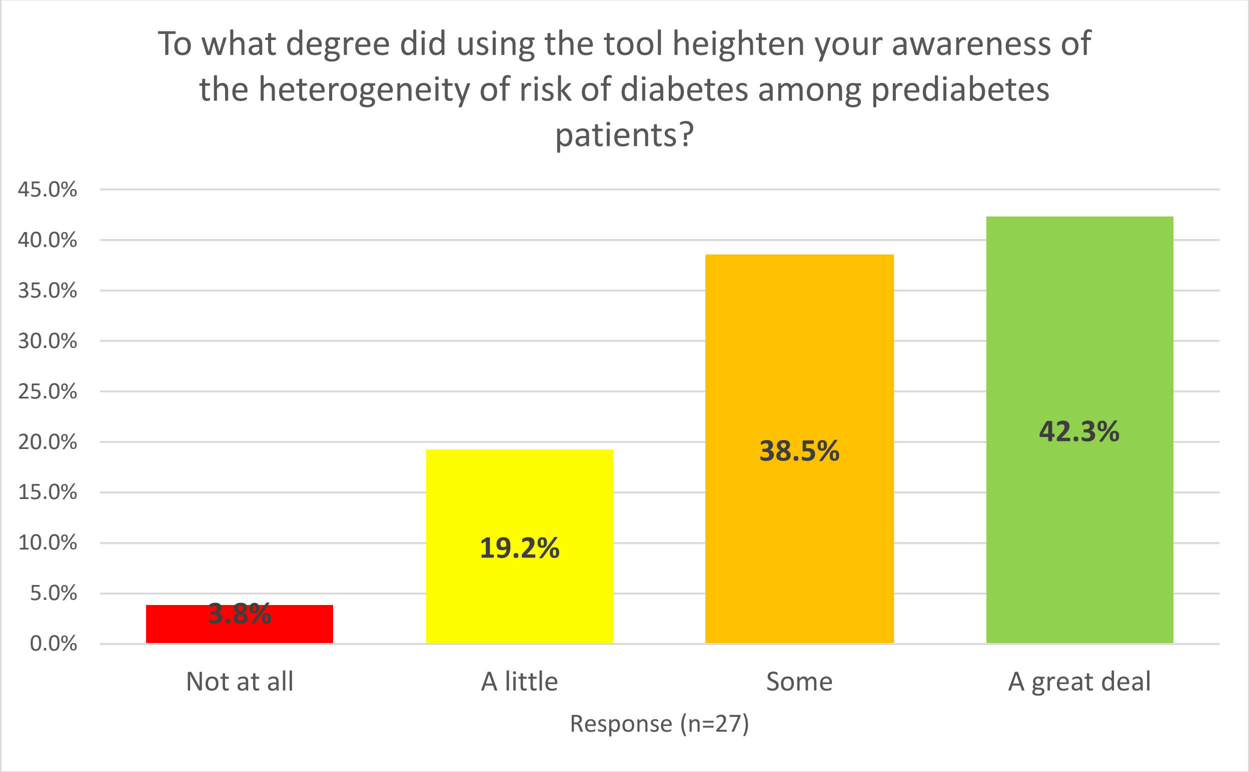
To what degree did using the tool heighten your awareness of the heterogeneity of risk of diabetes among prediabetes patients?
45.0%
40.0%
35.0%
30.0%
25.0%
20.0%
15.0%
10.0%
5.0%
0.0%
3.8%
19.2%
38.5%
42.3%
Not at all
A little
Some
A great deal
Response (n=27)

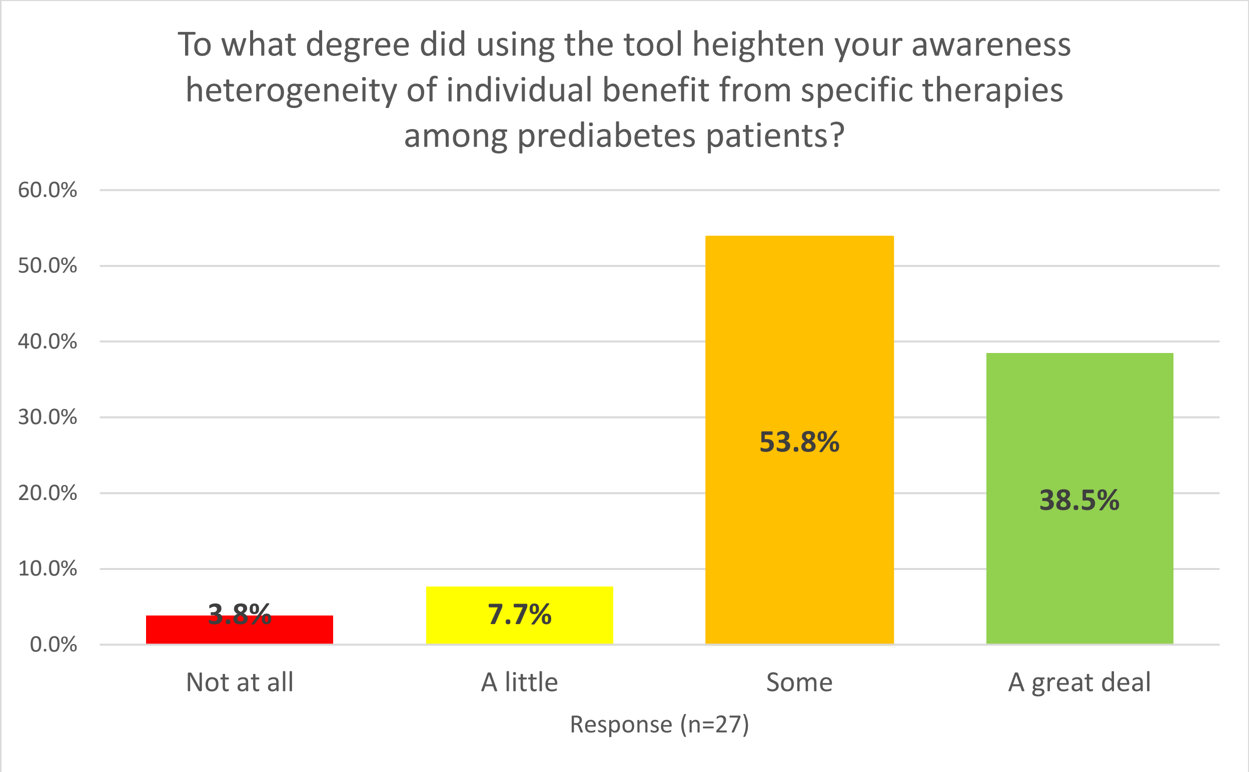
To what degree did using the tool heighten your awareness heterogeneity of individual benefit from specific therapies among prediabetes patients?
60.0%
50.0%
40.0%
30.0%
20.0%
10.0%
0.0%
3.8%
7.7%
53.8%
38.5%
Not at all
A little
Some
A great deal
Response (n=27)

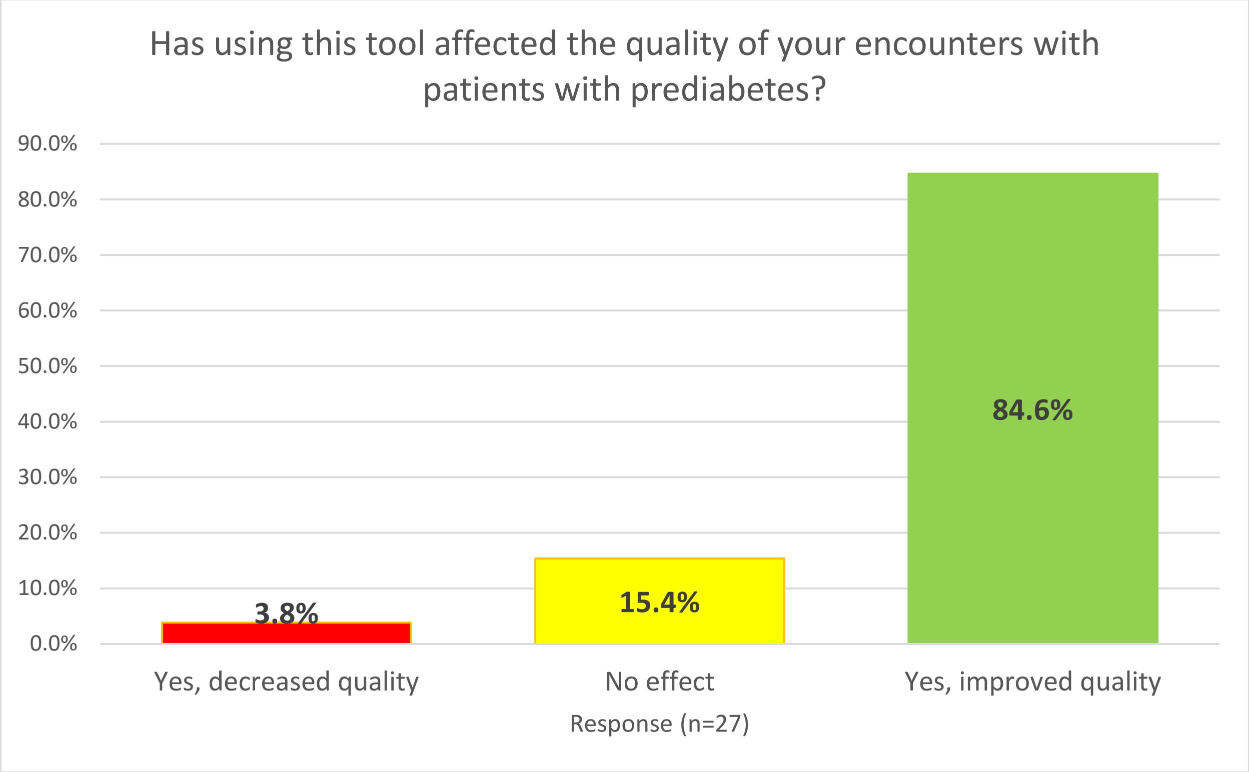
Has using this tool affected the quality of your encounters with patients with prediabetes?
90.0%
80.0%
70.0%
60.0%
50.0%
40.0%
30.0%
20.0%
10.0%
0.0%
3.8%
15.4%
84.6%
Yes, decreased quality
No effect
Yes, improved quality
Response (n=27)

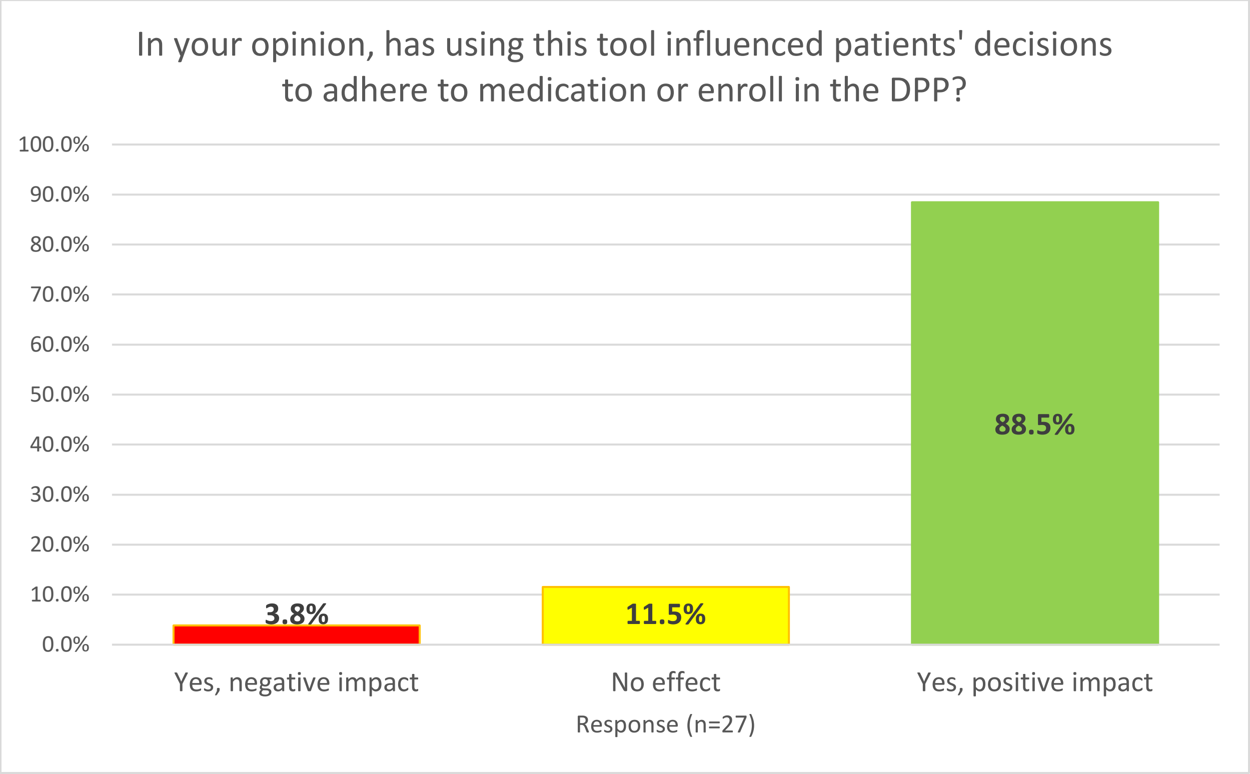
In your opinion, has using this tool influenced patients' decisions to adhere to medication or enroll in the DPP?
100.0%
90.0%
80.0%
70.0%
60.0%
50.0%
40.0%
30.0%
20.0%
10.0%
0.0%
3.8%
11.5%
88.5%
Yes, negative impact
No effect
Yes, positive impact
Response (n=27)

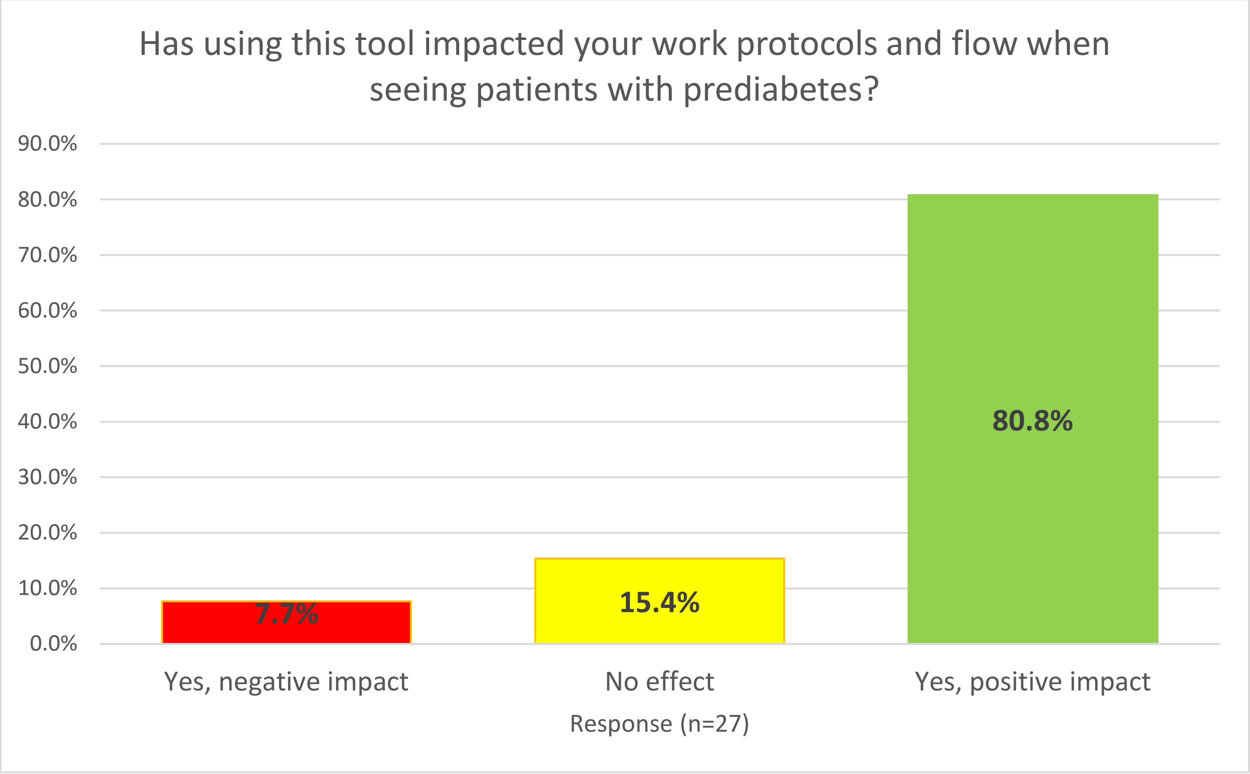
Has using this tool impacted your work protocols and flow when seeing patients with prediabetes?
90.0%
80.0%
70.0%
60.0%
50.0%
40.0%
30.0%
20.0%
10.0%
0.0%
7.7%
15.4%
80.8%
Yes, negative impact
No effect
Yes, positive impact
Response (n=27)

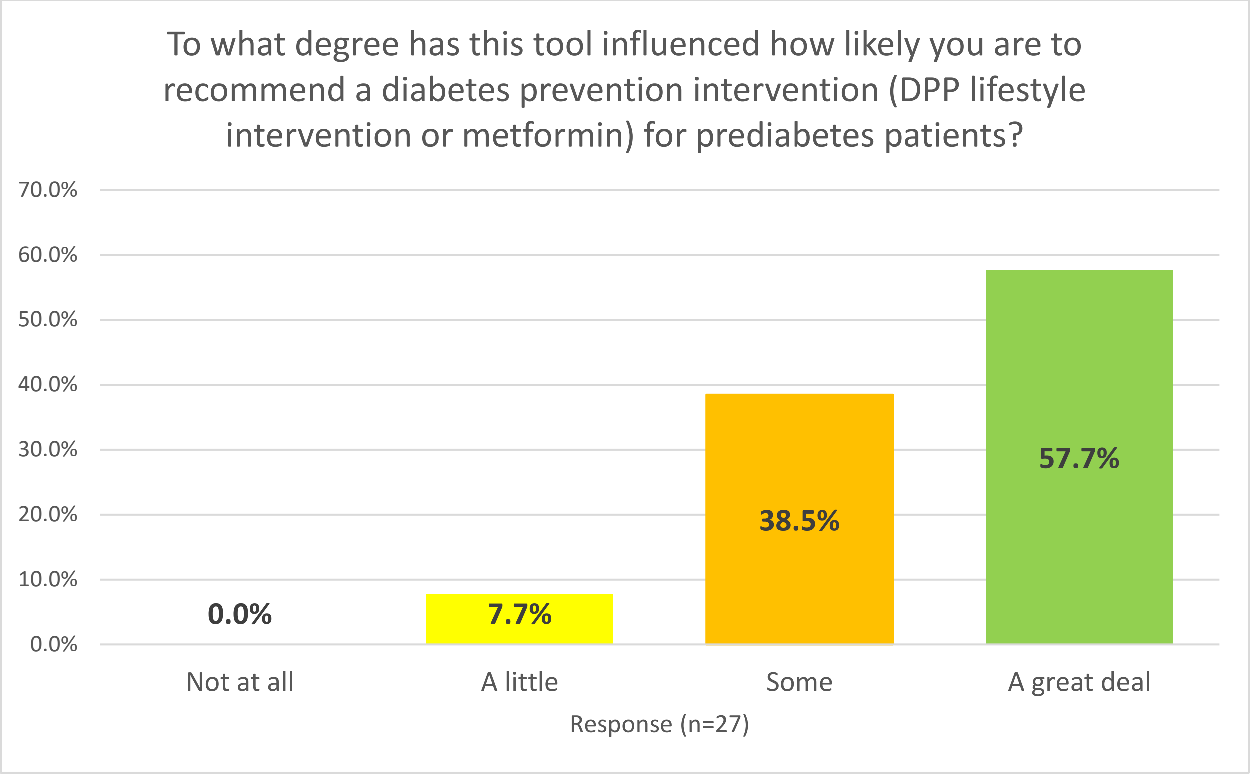
To what degree has this tool influenced how likely you are to recommend a diabetes prevention intervention (DPP lifestyle intervention or metformin) for prediabetes patients?
70.0%
60.0%
50.0%
40.0%
30.0%
20.0%
10.0%
0.0%
0.0%
7.7%
38.5%
57.7%
Not at all
A little
Some
A great deal
Response (n=27)

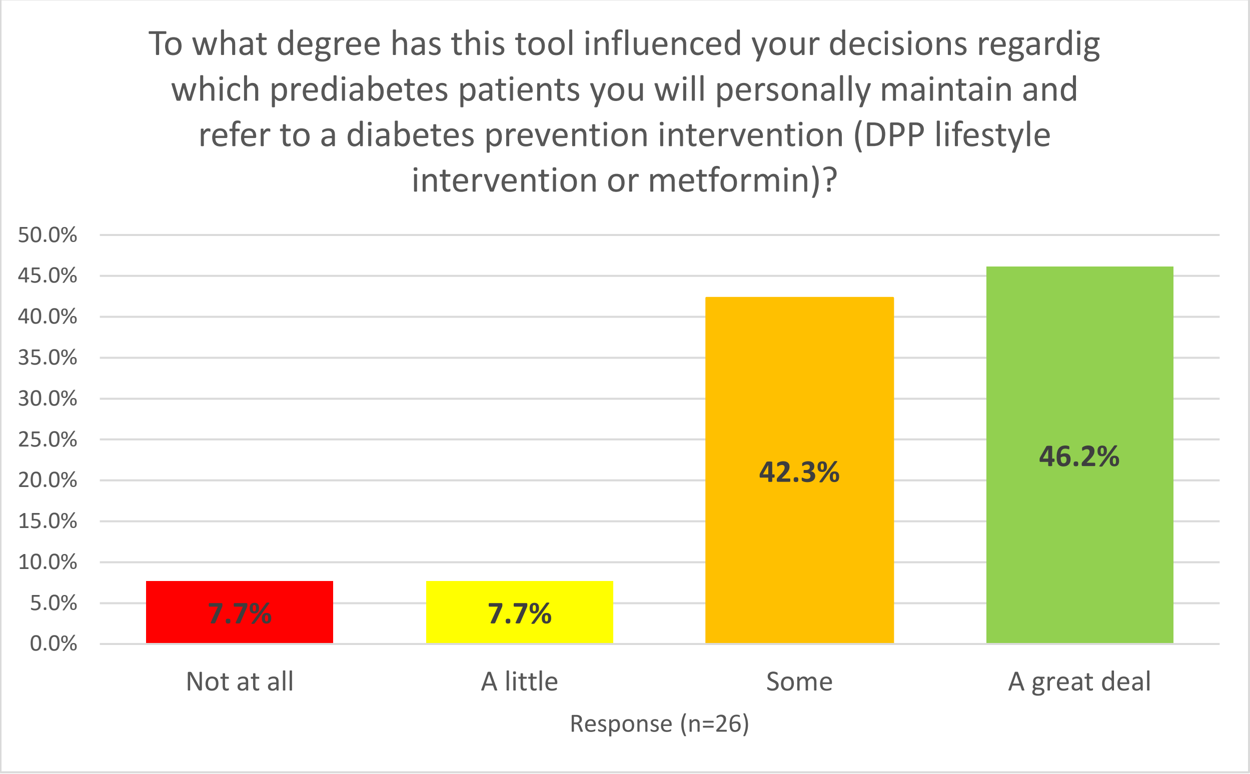
To what degree has this tool influenced your decisions regardig which prediabetes patients you will personally maintain and refer to a diabetes prevention intervention (DPP lifestyle intervention or metformin)?
50.0%
45.0%
40.0%
35.0%
30.0%
25.0%
20.0%
15.0%
10.0%
5.0%
0.0%
7.7%
7.7%
42.3%
46.2%
Not at all
A little
Some
A great deal
Response (n=26)
