## Additional File 3. Outcomes study additional tables and figures for "Effect of discussing personalized estimates of diabetes risk for people with prediabetes"

I. Definition of Progression to Diabetes

Progression to Diabetes based on claims data is being defined as follows:

1. Type 2 – two or more distinct claims with E11.xx diagnosis in primary or any secondary field, or
2. Rx claim(s) in the post-period for a total of 60 days or more of a diabetes drug from **Table A** and no type 1 diagnosis code on any claim in the post-period and a type 2 diagnosis code on at least one post-period claim, or
3. Rx claim(s) in the post-period for a total of 60 days or more of a diabetes drug from **Table B**, and a type 2 diagnosis code on at least one post-period claim.

**Table A**

| **STC Code** | **STC Description** |
|---|---|
| 0178 | Antihyperglycemic, insulin release stimulant type |
| 0179 | Antihyperglycemic, biguanide type |
| 7768 | Antihyperglycemic, alpha-glucosidase inhibitors |
| 7769 | Antihyperglycemic, thazolidinedione (PPARG Agonist) |
| A716 | Antihyperglycemic, amylin analog-type |
| B135 | Antihyperglycemic, thiazolidinedione-sulfonylurea |
| B137 | Antihyperglycemic, insulin-release stim. -Biguanide |
| B139 | Antihyperglycemic, thiazolidinedione and biguanide |
| B789 | Antihyperglycemic, DPP-4 inhibitors |
| C118 | Antihyperglycemic, DPP-4 inhibitor-biguanide combs |
| D549 | Antihyperglycemic, dopamine receptor agonists |
| E191 | Antihyperglycemic-glucocorticoid receptor blocker |
| E874 | DPP-4 Enzyme inhib - thiazolidinedione |
| F555 | Antihyperglycemic, SGLT2 inhibitor-biguanide combs |
| F883 | Antihyperglycemic, SGLT2 and DPP-4 inhibitor comb |

**Table B**

| STC Code | STC Description |
|---|---|
| 0179 | Antihyperglycemic, biguanide type |
| A771 | Antihyperglycemic, incretin mimetic (GLP-1 Receptor agonist) |
| E948 | Antihyperglycemic-sodium/glucose cotransporter (SGLT2) inhibitor |
| 0277 | Bromocriptine mesylate (Antiparkinsonism drug, other) |

II. Sample selection for outcomes analysis

A: Commercially insured patients

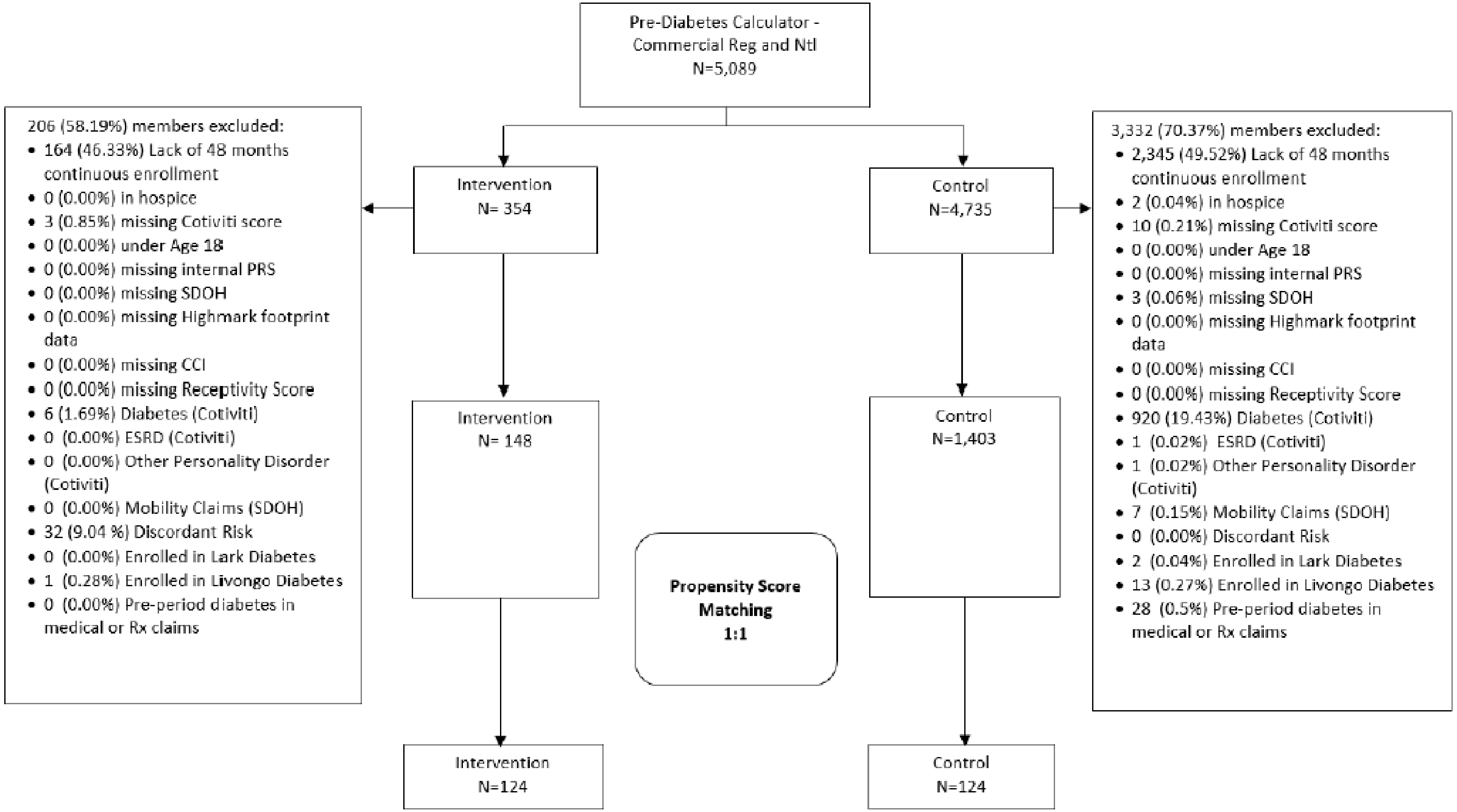

B: Medicare Advantage patients

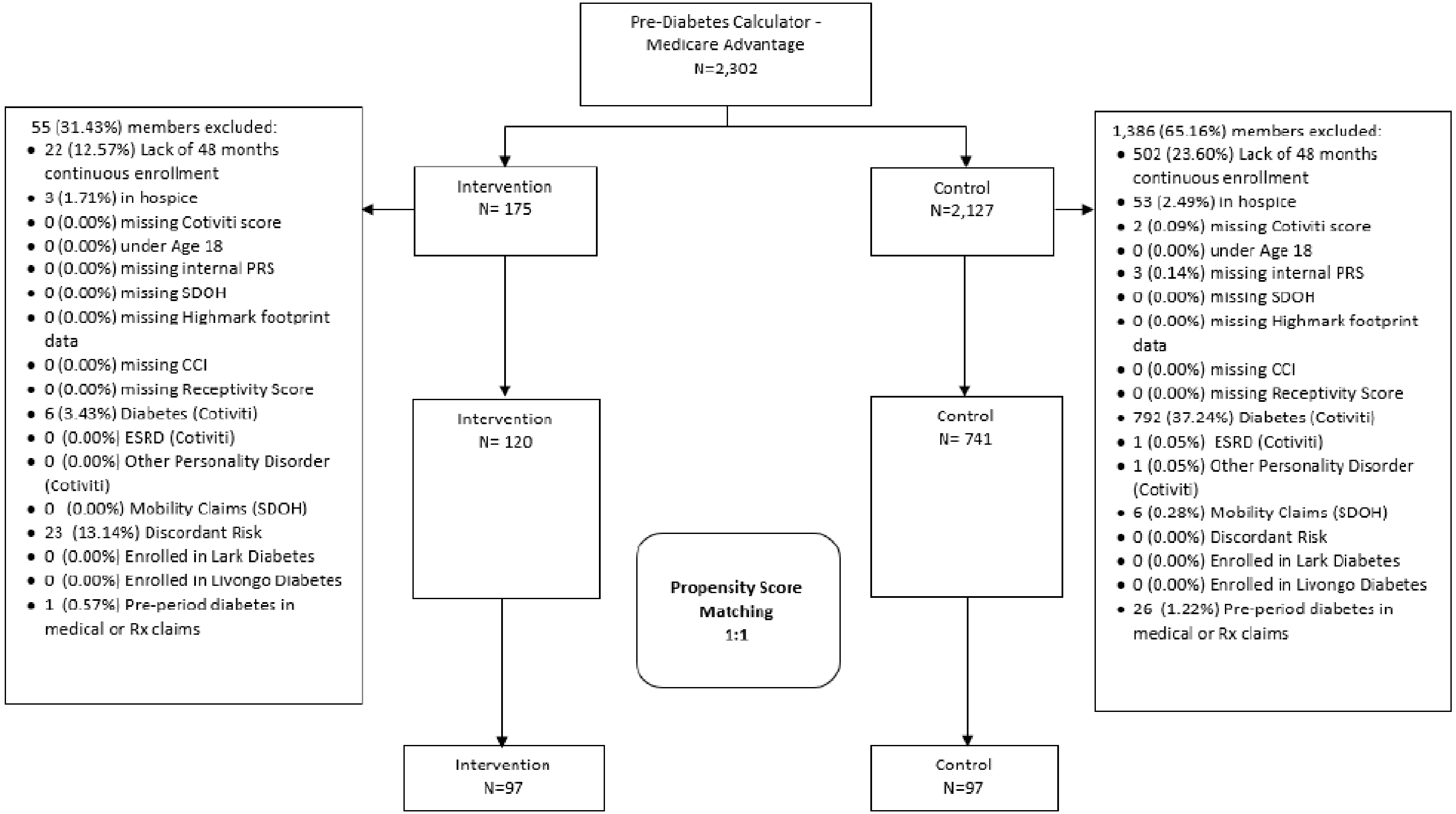
